# Infant respiratory syncytial virus and childhood asthma: a nationwide registry-based, sibling-controlled, and genome-wide association study

**DOI:** 10.64898/2026.07.17.26351934

**Authors:** Pekka Vartiainen, Hele Haapaniemi, Yunsung Lee, Maria C Magnus, Tuomo Hartonen, Kira Detrois, Essi Viippola, Matteo Ferro, Tarja Laitinen, FinnGen, Maiken A Madsen, Sisse R Ostrowski, Ole B Pedersen, Erik Sørensen, Christian Erikstrup, Tong Gong, Samuel Rhedin, Cecilia Lundholm, Giulia Dallagiacoma, Catarina Almqvist, Amanda M Egeskov-Cavling, Thea K Fischer, Anu Pasanen, Mika Rämet, Anna-Leena Vuorinen, Tero Hiekkalinna, Siri E Håberg, Per Magnus, Markus Perola, Astanand Jugessur, Andrea Ganna, Santtu Heinonen

## Abstract

**Background:** Early-life respiratory syncytial virus (RSV) infection is associated with childhood recurrent wheeze or asthma (RW/A), but causality and shared genetic liability remain unclear.

**Methods:** We combined Finnish nationwide registries and Nordic genetic cohorts. First, in 965 312 Finnish children born between 1998 and 2014, we defined severe RSV as RSV hospitalisation before age 1 year, and recurrent wheezing or asthma (RW/A) as inhaled medication reimbursement between ages 1 and 7 years, and compared medication and eosinophil trajectories by RSV history. Second, we assessed familial confounding in 527 776 full siblings and 15 667 RW/A- discordant pairs. Third, we performed a genome-wide association study (GWAS) of RSV susceptibility with meta-analysis across six Nordic cohorts (3 107 cases, 92 091 controls) and two-sample Mendelian randomisation (2SMR) using 155 asthma-associated variants.

**Findings:** RSV-associated RW/A showed higher inhaled medication use at ages 1–2 years but lower use after age 5, and lower mean blood eosinophils (0·341 vs 0·398×10□/L; p=0·008) than RW/A without RSV hospitalisation. In RW/A-discordant sibling pairs, RSV hospitalisation was associated with RW/A (OR 2·8; 95% CI 2·4–3·2), while unaffected siblings also had elevated RW/A prevalence. GWAS identified an RSV association at *APBB1IP* (rs787036; β=0·209; p=8·80×10^-^□). 2SMR provided no evidence that asthma genetic liability influenced RSV susceptibility.

**Interpretation:** The RSV–asthma association is unlikely to be explained by shared genetic or environmental factors, and RSV-associated RW/A shows a distinct trajectory. These findings help prioritise long-term outcomes for RSV prevention trials and monitoring.

**Funding:** Päivikki and Sakari Sohlberg Foundation, Foundation for Pediatric Research, Sigrid Jusélius Foundation, Orion Research Foundation, the Research Council of Norway.

## Introduction

Asthma is the most common chronic disease of childhood, affecting 10% of children globally, and up to 40% of cases may be preventable by modifying environmental risk factors.^1^ Respiratory syncytial virus (RSV) infection has long been associated with recurrent wheezing and asthma later in childhood, particularly when RSV causes bronchiolitis or requires hospitalisation in infancy.^2,3^ However, whether RSV contributes causally to asthma development or instead marks an underlying susceptibility remains unresolved,^2^ and previous studies with passive RSV immunisation have not demonstrated a reduction in childhood asthma.^4,5^

Asthma encompasses multiple disease endotypes with distinct clinical and biological characteristics. In young children, recurrent viral-triggered wheeze can resemble asthma clinically but often improves or resolves before school age. Cohort studies suggest that asthma or recurrent wheeze following early-life RSV is often non-atopic and concentrated in the first years of life.^3,6^ The extent to which RSV and asthma share genetic liability is also poorly understood. Despite advances in asthma genetics,^7^ genome-wide association studies (GWAS) of RSV infection have been underpowered to identify significant loci.^8–12^ While certain asthma-associated variants have been linked to rhinovirus-induced wheezing,^13,14^ similar associations with RSV are not established.

Recently, a maternal RSV vaccine and two long-acting monoclonal antibodies, nirsevimab and clesrovimab, have become available to prevent RSV disease in infants. Although these interventions are highly effective against severe RSV disease, they do not prevent the infection.^15^ As RSV immunisation programmes are implemented, it will be crucial to determine whether later asthma risk is associated with RSV infection broadly, with disease severity, or with the underlying susceptibility of children who develop severe RSV disease.

## Aims

We aimed to characterise how early-life severe RSV infection relates to asthma from clinical, familial, and genetic perspectives by (i) comparing inhaled medication use and eosinophil trajectories in children with and without infant RSV hospitalisation; (ii) evaluating shared familial contributions using discordant full-sibling analyses; (iii) performing an RSV GWAS meta-analysis to identify susceptibility loci; and (iv) using two-sample Mendelian randomisation to test whether asthma genetic liability influences RSV susceptibility.

## Methods

### Study design

We used three complementary designs: (i) nationwide registry linkage to examine longitudinal medication and laboratory measures; (ii) discordant full-sibling analyses to address confounding by shared genetics and familial factors; and (iii) genome-wide analyses of RSV across multiple cohorts, including GWAS meta-analysis with replication, and two-sample Mendelian randomisation to assess directionality. Reporting followed the STREGA and STROBE-MR guidelines.^16,17^

### Registry-based cohorts and variables

#### Population-based cohort and full sibling sub-cohort

We conducted registry-based analyses in a nationwide cohort of 965 312 children born in Finland between 1 Jan 1998 and 31 Dec 2014, excluding children who died (n=3 548) or emigrated (n=3 928) before age seven years. Registry follow-up ended on 31 Dec 2021, providing complete follow-up for the primary asthma outcome through age seven. For descriptive purposes, we followed drug purchases and blood eosinophil measurements from age one until the age of 12 years.

For within-family analyses, we identified 527 776 children forming 263 888 full-sibling pairs (first- and second-born), born to the same parents ≤6 years apart within the same birth years. Twins (n=28 328) and children with missing father’s identifier (n=19 814) were excluded from sibling analyses (**Figure 1**). For additional cohort and methodological details, see **Supplementary Material**.

**Figure 1.**
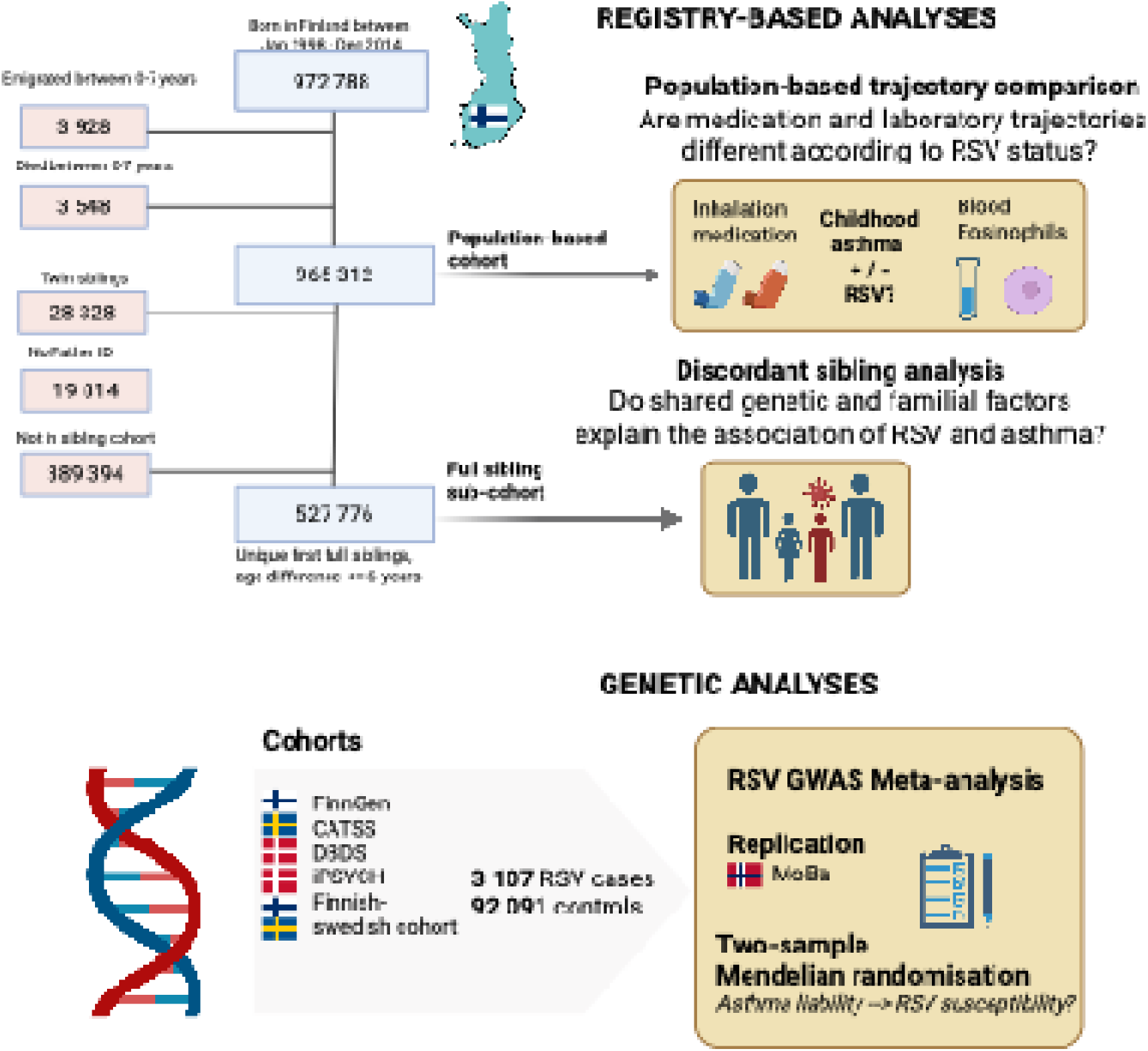
Overview of the study design and cohorts. The flowchart in top left shows numbers and exclusions at each step of the registry-based cohort definitions. RSV = Respiratory Syncytial Virus. GWAS = Genome-Wide Association Study. FinnGen = Finnish biobank collaboration project. CATSS = The Child and Adolescent Twin Study in Sweden. DBDS = Danish Blood Donor Study iPSYCH = Previous publication from Danish iPSYCH cohort.^10^ Finnish-Swedish cohort: previous publication by Pasanen et al.^8^ MoBa = Norwegian Mother, Father and Child Cohort study

#### Data sources

In the registry-based analyses, we used data from the Medical Birth Registry, Hospital Discharge Registry, National Drug Purchase Registry and the Kanta laboratory archive in Finland. These registries and the preprocessing are described in more detail in the Supplementary Material and the cohort profile paper.^18^ All prescription drug purchases reimbursed by the national health insurance, including all inhaled medication purchases and reimbursement codes, are recorded in the National Drug Purchase Registry. The Kanta laboratory database contains all laboratory measurements performed in clinical practice across public and private healthcare providers. Data collection started in 2014 with healthcare providers joining the system gradually. Full or near-full coverage was reached in 2019 when all private healthcare providers were also required to provide data.

#### Variable definitions

##### RSV hospitalisation

In the registry-based analyses, we defined severe RSV as inpatient RSV hospitalisation during the first year of life, using the International Classification of Diseases, 10th Revision (ICD-10) codes J21.0, J12.1 and J20.5. In Finnish registry data, the RSV diagnoses correspond to microbiological confirmation.^19^ We restricted RSV to hospitalisations at ≤1 year to limit surveillance bias (older children with emerging asthma could be tested more often) and age-related differences in testing beyond infancy. Confining exposure to the first year of life also preserved temporal ordering with later wheezing/asthma, reducing concerns about reverse causation.

##### Childhood recurrent wheezing/asthma

As the primary definition of childhood recurrent wheezing/asthma (RW/A), we used a special reimbursement status (code 203) for inhaled medication granted by Kela – the Social Insurance Institution of Finland – between ages one and seven years. Entitlement requires a submitted application with a physician’s statement confirming age-specific diagnostic criteria for asthma and documenting adequate follow-up. In children younger than 3 years, eligibility requires recurrent physician-confirmed wheezing episodes. The number of wheezing episodes required depends on the severity, frequency, and presence of additional risk factors (such as allergies or positive family history) ranging from 2 episodes (if exceptionally severe or frequent), to 3 or 4 episodes (with and without risk factors, respectively) within one year. Importantly, early life RSV bronchiolitis should not be considered when counting the recurrent wheezing episodes. In addition, regular anti-inflammatory inhaled medication therapy is required during the preceding 6 months. In children aged 4-6 years, signs of reversible obstruction in lung function tests are also criteria. In descriptive analyses, we further grouped children with RW/A into RSV-RW/A (those with recurrent wheezing/asthma and prior RSV hospitalisation during the first year of life) and non-RSV-RW/A (those with recurrent wheezing/asthma and no RSV hospitalisation).

##### Drug purchase event definitions

We examined drug purchase data using Anatomical Therapeutic Chemical (ATC) codes from the National Drug Purchase Registry. To identify inhaled medication purchases, we used the 5-character ATC codes R03AC for bronchodilators, and R03BA and R03AK for inhaled corticosteroids (ICS). Among sibling pairs discordant for RSV hospitalisation, we also examined rates of commonly-used antibiotics for bacterial respiratory tract infections (amoxicillin, ATC code J01CA04 and amoxicillin-clavulanate, J01CR02) as an additional outcome.

### Genetic cohorts and phenotypes

#### Genotyped cohorts

The GWAS meta-analysis of RSV susceptibility incorporated the studies and datasets as described below. For more details, see the **Supplementary Material.**

The biobank-based **FinnGen** study includes data from 520 000 Finnish individuals linked to extensive national registry data.^20^ The Child and Adolescent Twin Study in Sweden (CATSS) is an ongoing, population-based longitudinal cohort within the Swedish Twin Registry, targeting all twins born in Sweden since 1 July, 1992.^21^ The Danish Blood Donor Study (DBDS) is an ongoing cohort and biobank comprising over 135 000 genotyped participants.^22^ In addition to these cohorts, we incorporated summary statistics from two previously published GWAS of early-life RSV, from the Danish iPSYCH cohort and from a Finnish-Swedish cohort.^8,10^ Analyses were restricted to children born in 1995 or later, when microbiological RSV testing for infants became routine practice. We then sought to replicate our findings in the Norwegian Mother, Father and Child Cohort Study MoBa,^23^ a genotyped and nationwide pregnancy cohort in Norway between 1998 and 2008, using available questionnaire-based information on respiratory outcomes. In MoBa, out of 111 921 enrolled pregnancies, 63 341 genotyped children with non-missing 6-month questionnaire data were included in the replication.

#### GWAS case and control definitions

For the GWAS meta-analysis, the cohorts adhered to a shared analysis plan containing the phenotype definitions and quality control (QC) steps, shown in revised form in the Supplementary material and summarised below.

The GWAS phenotype was any RSV-specific lower respiratory tract infection (LRTI) diagnosis, either from inpatient or outpatient care, during the first 2 years of life and defined using ICD-10 codes J21.0, J20.5 or J12.1. We chose a broad phenotype to harmonise with previous studies and maximise the number of cases for the GWAS. An exception was the previous iPSYCH study,^10^ where the outcome was RSV hospitalisation before 5 years of age. We deemed this to be acceptable because only a small subset of RSV hospitalisations occurred after the second year of life.^10^ As a FinnGen-specific sensitivity analysis, we also defined RSV cases using inpatient diagnoses only.

For GWAS replication in the MoBa cohort, parental report of any LRTI by 6 months of age (positive questionnaire response for RSV/bronchitis/pneumonia) was used as the outcome. This measure was considered the closest available proxy to an RSV-specific diagnosis-based phenotype, as most LRTI episodes in early infancy are attributable to RSV and complete diagnosis data were not available.

For case and control numbers in each cohort, see Supplementary Table 3, and further details on cohorts, phenotypes and QC are found in the Supplementary material.

### Statistical analyses

#### Registry-based analyses

In the population cohort, we performed two descriptive analyses: (i) annual inhaled medication purchase rates and (ii) blood eosinophil trajectories, stratified by infant RSV hospitalisation and RW/A. Medication trajectories were summarised as mean purchases per person-year of age with 95% confidence intervals (CI). For eosinophils, we extracted absolute eosinophil counts (10 /L) from the Kanta laboratory archive, and derived each child’s median eosinophil value for each year of age with available data (one observation per child-year). Group-level trajectories across the four RSV–RW/A strata were visualised using generalised additive models (GAMs) fitted to these yearly medians; as sensitivity analyses, generalised additive mixed models (GAMMs) with subject-specific random intercepts were fitted among children with asthma, with and without RSV. Group means were compared using independent-samples t-tests.

In the Finnish full-sibling sub-cohort, we compared RW/A prevalence by age 1-7 years in RSV-concordant and RSV-discordant full-sibling pairs, with sensitivity analyses by birth order and parental asthma. We then used conditional logistic regression with RW/A as the outcome to control for shared familial factors, adjusting for sex, gestational age, birth month, birth year, caesarean delivery, maternal smoking during pregnancy, and birth order (A directed acyclic graph (DAG) is shown in Supplementary Figure 2).

All registry-based analyses were performed in R (v4.2.0).

##### Missing or incomplete data

Missingness in Finnish registries is rare; many events of interest (such as hospitalisations or prescription drug purchases) are automatically and comprehensively recorded. Father’s identifier was missing for 19 814 (2·1%) children, who were therefore excluded from sibling-based analyses. There were missing values in gestational age (0·16%) and maternal smoking during pregnancy (2·4%), and those cases were excluded from the conditional logistic regression. For trajectory visualisations, follow-up was extended to age 12 for descriptive purposes; children born after 2009 contributed available observations, and their follow-up beyond age 7 was incomplete. See the **Supplementary Material** for more details.

#### Genetic analyses

Each participating cohort in the GWAS meta-analysis first performed their GWAS of the RSV phenotype using a standard quality control (QC) protocol. In summary, per-sample QC required a call rate ≥0·98, heterozygosity (FHET) within ±0·20, and exclusion of individuals with sex discrepancies. Per-variant QC required a call rate ≥0·95, differential missingness ≤0·02, and Hardy-Weinberg equilibrium (HWE) p≥1×10_-_□ in controls and ≥1×10⁻¹□in cases. For more details, see the **Supplementary Material** and the original cohort descriptions.

We combined cohort-specific GWAS summary statistics with fixed-effects inverse-variance meta-analysis. This is the standard and more powerful approach for GWAS discovery meta-analysis assuming shared allelic effect across cohorts, in contrast to random effects meta-analysis where the effect is assumed to vary across cohorts. We used METAL software^24^ for the GWAS meta-analysis, restricting to common variants (minor allele frequency > 1%). We evaluated between-cohort heterogeneity with Cochran’s Q and *I*^2^. In the external replication cohort (MoBa), we tested associations of the lead single nucleotide polymorphism (SNP) and an RSV polygenic score (derived with the PRS-CS method)^25^ with the questionnaire-based LRTI outcome using additive logistic regression adjusted for sex, birth year, imputation batch, and principal components 1–20.

As a FinnGen-specific sensitivity analysis, we repeated the RSV GWAS using an inpatient-only case definition and compared it with the primary FinnGen GWAS (including both in- and outpatient RSV cases). We assessed concordance with Linkage Disequilibrium Score Regression (LDSC) and by comparing effect estimates for the genome-wide significant variants from the main meta-analysis.

##### Two-sample Mendelian Randomisation (MR)

We selected genome-wide significant (p<5×10^-^□) asthma variants from the published asthma GWAS,^7^ clumped these variants for independence using 1000 Genomes Phase 3 European reference (10 Mb, r²<0·001), and retained the lead SNP per locus. Two-sample MR was performed by harmonising these instruments with the RSV GWAS meta-analysis summary statistics to test whether genetic liability to asthma causally influences RSV susceptibility. As a sensitivity analysis, we repeated the MR by using only childhood asthma -specific variants.^7^

## Ethics approval

The Finnish registry-based analyses were conducted within the FinRekisterit2025 project, coordinated by the Finnish Institute for Health and Welfare (THL) and approved under decisions THL/5757/6.02.00/2025, THL/2844/6.02.00/2023, and THL/6366/6.02.00/2024. Genetic analyses in contributing genotyped cohorts were conducted under the relevant cohort-specific ethics approvals, consent procedures, and data-use permissions. Further details are provided in the Supplementary Material.

## Results

### RSV-associated recurrent wheezing/asthma shows early inhaled medication use and lower eosinophils

In the population-based cohort (N = 965 312), 14 117 children (1·5%) experienced RSV hospitalisation during the first year of life, and 35 343 (3·7%) received a special reimbursement for inhaled medications due to recurrent wheezing/asthma between ages one and seven years (RW/A). See **Supplementary Table 1** for background characteristics. Overall, 33 479 children (3·5%) had RW/A but no RSV hospitalisation (non-RSV-RW/A) and 1 864 (0·2%) had both RSV hospitalisation and RW/A (RSV-RW/A).

The crude risk ratio of RW/A following RSV hospitalisation was 3·71 (95% CI: 3·56–3·87). Children with RSV-RW/A had an earlier onset (mean age 2·27 years, 95% CI 2·19–2·34) than children with non-RSV-RW/A (3·35 years, 95% CI 3·33–3·37). Children with RSV-RW/A used more inhaled steroids between ages 1 and 2 years (2·62 vs 1·47 purchases per person, mean difference (95% CI) 1·15 (1·04–1·27), p = 3·3e-75), and less at 5-6 years (1·65 vs 2·32 purchases, mean difference 0·68, 95% CI 0·57–0·79, p = 1·55×10^-34^) and afterwards, compared to non-RSV-RW/A, indicating earlier discontinuation or less need for inhaled steroids at the population level **(Figure 2)**. The trend was similar for bronchodilators, but the differences between the RSV- and non-RSV-RW/A groups were smaller (**Figure 2**).

**Figure 2:**
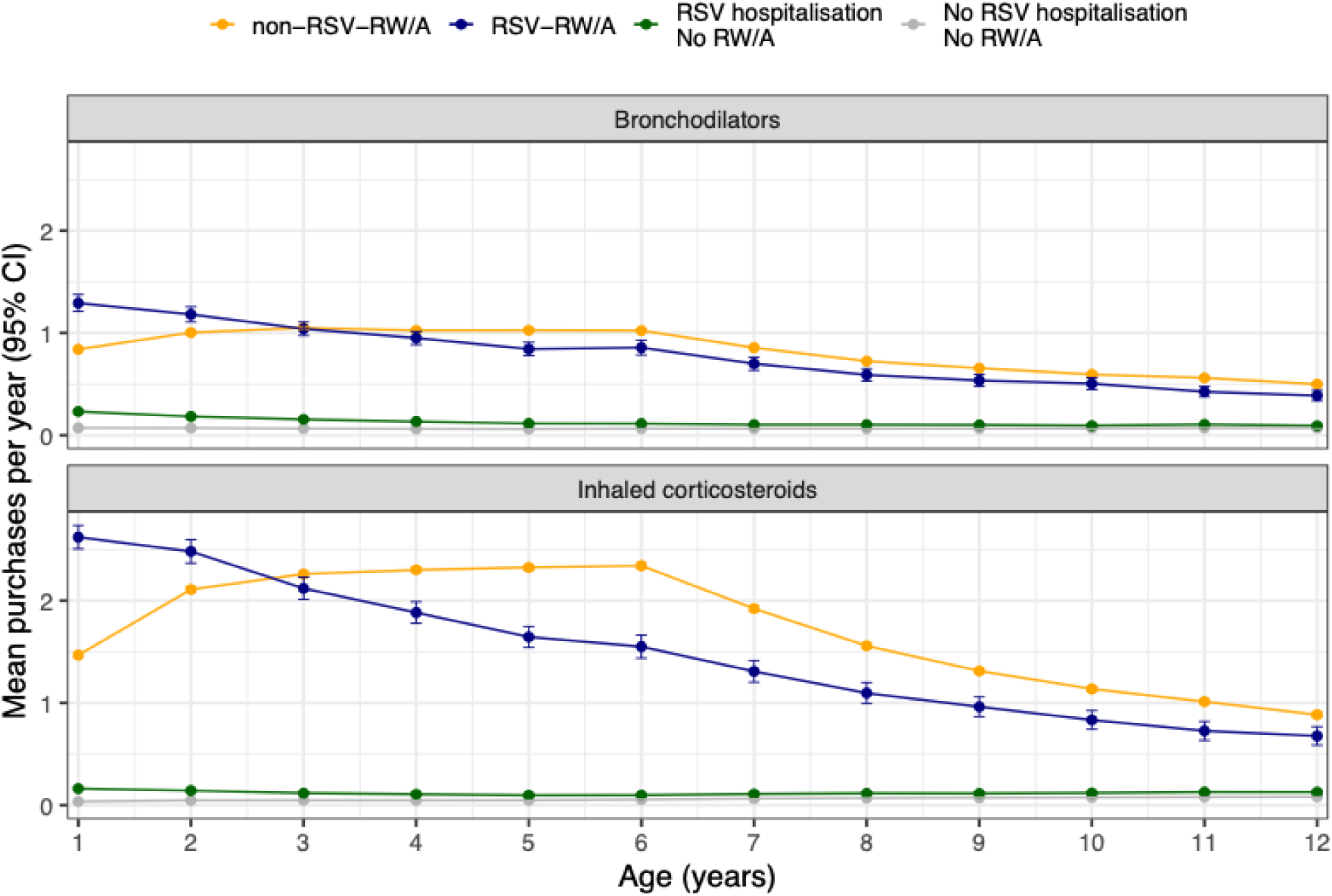
Mean annual purchases of bronchodilators and inhaled corticosteroids (ICS) from age one to age 12 years, stratified by infant RSV hospitalisation (by age ≤12 months) and childhood-onset recurrent wheezing/asthma (RW/A). Points show mean purchases per person-year and error bars indicate the 95% confidence intervals. RW/A was defined by special reimbursement for inhaled medication between ages 1–7 years. RSV-RW/A denotes children with both infant RSV hospitalisation and RW/A; non-RSV-RW/A denotes RW/A without RSV hospitalisation. Follow-up was censored at 31 Dec 2021 (children contributed data up to their last observed age). Children with purchases of bronchodilator nebuliser solutions (n=4 868) or budesonide nebuliser solutions (n=408) were excluded, as nebuliser use likely reflects a clinically distinct subgroup with different treatment patterns and severe comorbidity profiles. For example, children with RSV-RW/A had a statistically significant higher purchase rate in early life (ages 1–2 years), but lower purchase rates after age 5, particularly for ICS, compared with children with asthma but no RSV hospitalisation. The difference in ICS purchases for RSV-associated RW/A and RW/A without RSV was 2·22 vs 0·89 (p-value for difference 1·3×10^-99^) at age 1, and 1·75 vs 2·27 (p-value for difference 2·25×10^-25^) at age 5.

There were 77 947 children with at least one eosinophil measurement recorded between ages 1 and 12 years. Children with RSV-RW/A had, on average, lower median eosinophil levels than those with non-RSV-RW/A (0·341 vs 0·398×10^9^ cells/L, p = 0·008), and eosinophil trajectories indicated higher eosinophil counts between ages 3–8 years among non-RSV-RW/A **(Figure 3; Supplementary Figure 1)**.

**Figure 3.**
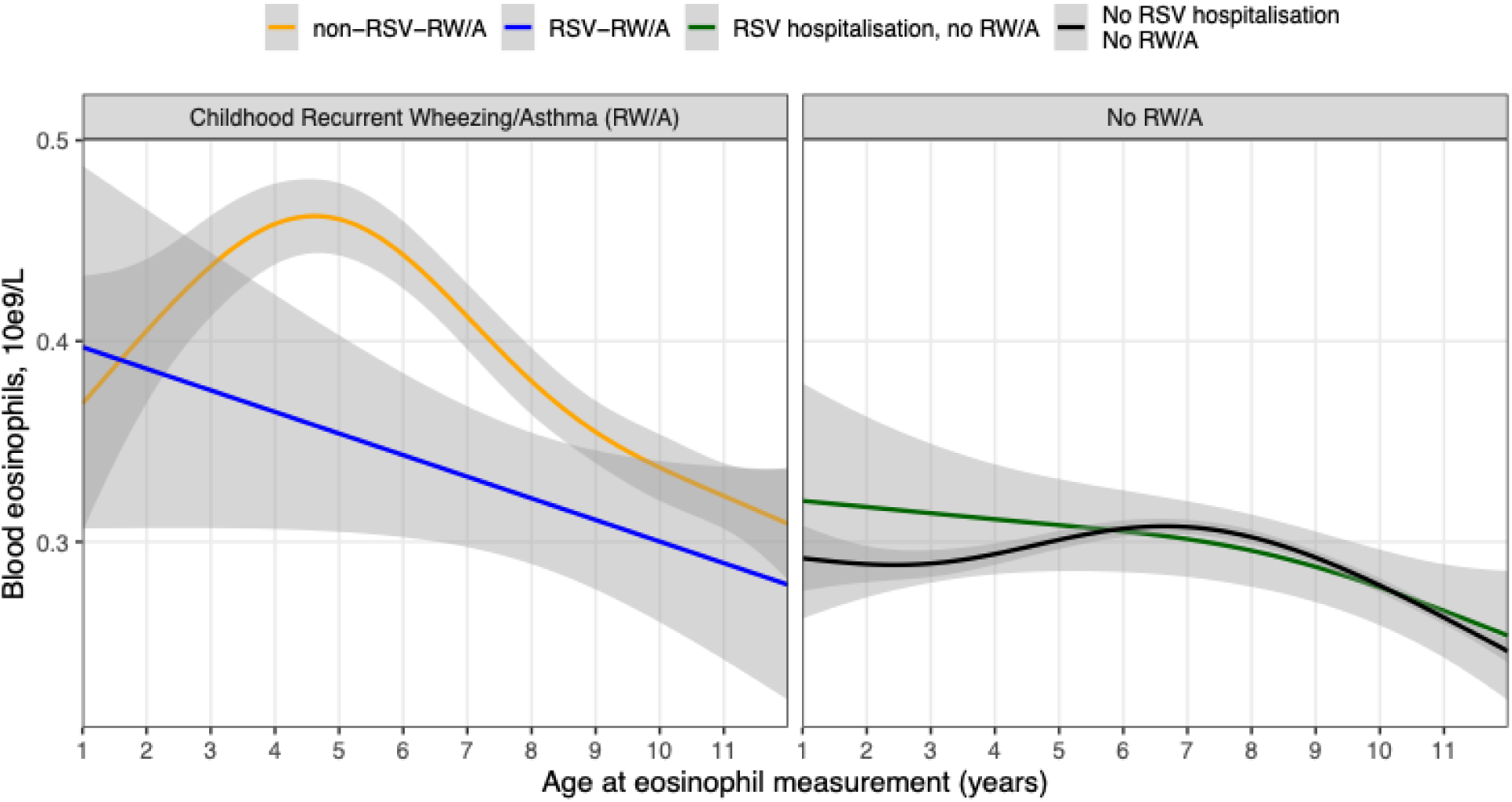
Comparison of eosinophil measurements stratified by early-life RSV hospitalisation and childhood-onset recurrent wheezing/asthma (RW/A). From each child, we extracted the median eosinophil value for each year of age, starting from age one. The curves show generalised additive models (GAM) fitted to these median values as a function of age, stratified into four groups according to RSV history and RW/A. The ribbons show the 95% confidence intervals, derived from the standard errors of the fitted GAM. Children with RSV-RW/A had lower eosinophil levels between ages 3 and 8 years than children with non-RSV-RW/A. Sensitivity analysis using a generalised additive mixed model (GAMM) showed a similar trend of lower eosinophils of RSV-RW/A compared to non-RSV-RW/A, but with smaller difference between the groups (**Supplementary** Figure 1). The analysis includes 97 758 eosinophil measurements from 71 932 children without RSV hospitalisation and without asthma; 6 588 measurements from 4 367 children with non-RSV-RW/A; 2 127 from 1 400 children with RSV hospitalisation but no asthma; and 402 measurements from 248 children with RSV- RW/A.

### Sibling comparisons support both shared familial susceptibility and an RSV-associated excess RW/A risk

To account for shared genetic, familial, and environmental confounding, we examined RW/A prevalence by 7 years of age in full-sibling pairs stratified by RSV hospitalisation during the first year of life. In a conditional logistic regression model adjusted for non-shared covariates among 15 667 sibling pairs discordant for RW/A, RSV hospitalization during the first year of life was strongly associated with RW/A (odds ratio (OR)=2·76, 95% CI: 2·43–3·15; p =1·62×10^-53^) (**Supplementary Table 2)**.

The pattern of higher RW/A rates in RSV-affected siblings remained after accounting for birth order (***Figure 4a***) and under different permutations of parental asthma status (**Figure 4b**). Notably, the unaffected sibling in RSV-discordant pairs also had around twice the asthma prevalence compared to siblings in whom neither had prior RSV hospitalisation **(Figure 4a)**. Importantly, the rates of antibiotic purchases were very similar between both siblings within RSV-discordant sibling pairs **(Supplementary Figure 3)**.

**Figure 4.**
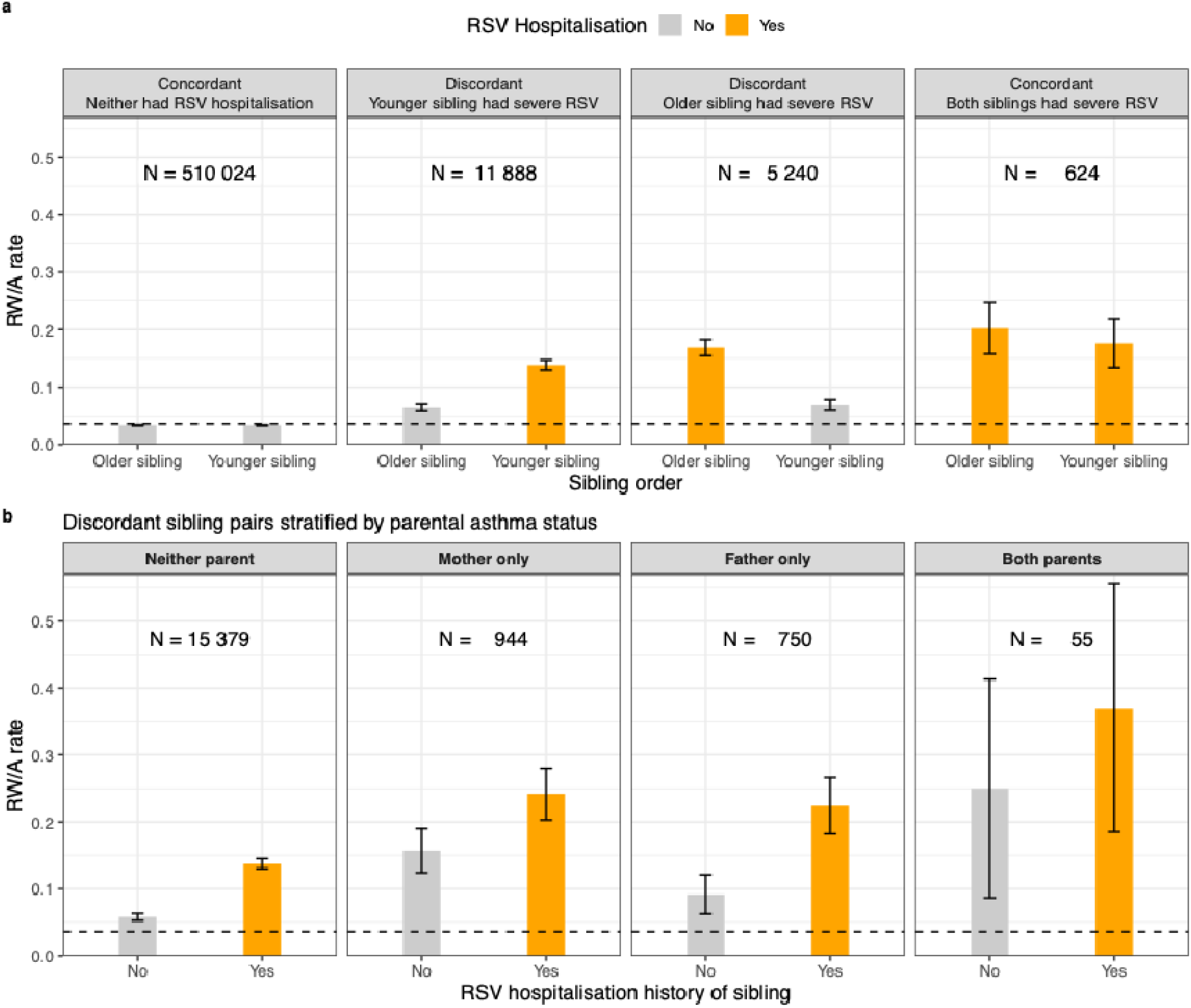
**Panel A** shows the rate of childhood-onset recurrent wheezing/asthma (RW/A, defined as special reimbursement for asthma medication between 1 and 7 years) in full-sibling pairs with an age difference of six years or less, stratified by RSV hospitalisation during the first year of life. The dashed line indicates the overall prevalence of the RW/A reimbursement by age 7 in the full population. In RSV-discordant sibling pairs, the asthma rate is approximately 2-3-fold higher in the RSV-affected sibling compared to the unaffected sibling, but the unaffected siblings in discordant pairs have roughly twofold higher asthma rates than siblings in pairs in which neither child has RSV hospitalisation. **Panel B** shows the prevalence of childhood recurrent wheezing/asthma reimbursement examined in RSV-discordant full-sibling pairs (i.e., first and second children of the same parents, where one sibling was hospitalised for RSV and the other was not), stratified by parental asthma status. Asthma in both, parents and children, is defined by the special reimbursement code for inhaled medications; for children this is assessed between ages 1–7 years, and for parents, before the birth of the child. Across all parental asthma strata, RSV-affected siblings exhibit higher recurrent wheezing/asthma prevalence than their non-affected siblings.

### GWAS meta-analysis of RSV revealed an association with variants in the *APBB1IP* gene

Our GWAS meta-analysis of early-life RSV infection, including 3 107 RSV cases and 92 091 controls, identified a single genome-wide significant locus with rs787036 as the lead SNP (chr10:26,505,176 A>G; GRCh38; p = 8·8×10^-^□) *–* an intronic variant in *APBB1IP* (***Supplementary Figure 4, locus zoom plot***). There was no evidence of heterogeneity between cohorts (Cochran’s Q = 3·05 with p = 0·69 and *I^2^* = 0 for the lead SNP rs787036). Genome-wide significant variants are listed in **Supplementary Table 4** and corresponding Q-Q plot is shown in **Supplementary Figure 5**. In the MoBa replication cohort (N = 63 341), the lead SNP rs787036 and an RSV polygenic score (RSV-PGS) were both associated with the questionnaire-based LRTI outcome by 6 months of age (**Figure 5b**). We found no clear pleiotropic effects of rs787036, as this variant was not associated with other diseases or with gene or protein expression in publicly available phenome-wide resources from FinnGen, UK Biobank, Open Targets, or Biobank Japan. SNP-based heritability in the RSV-GWAS was too low to support reliable genetic correlation estimates from Linkage Disequilibrium Score Regression (LDSC).

**Figure 5.**
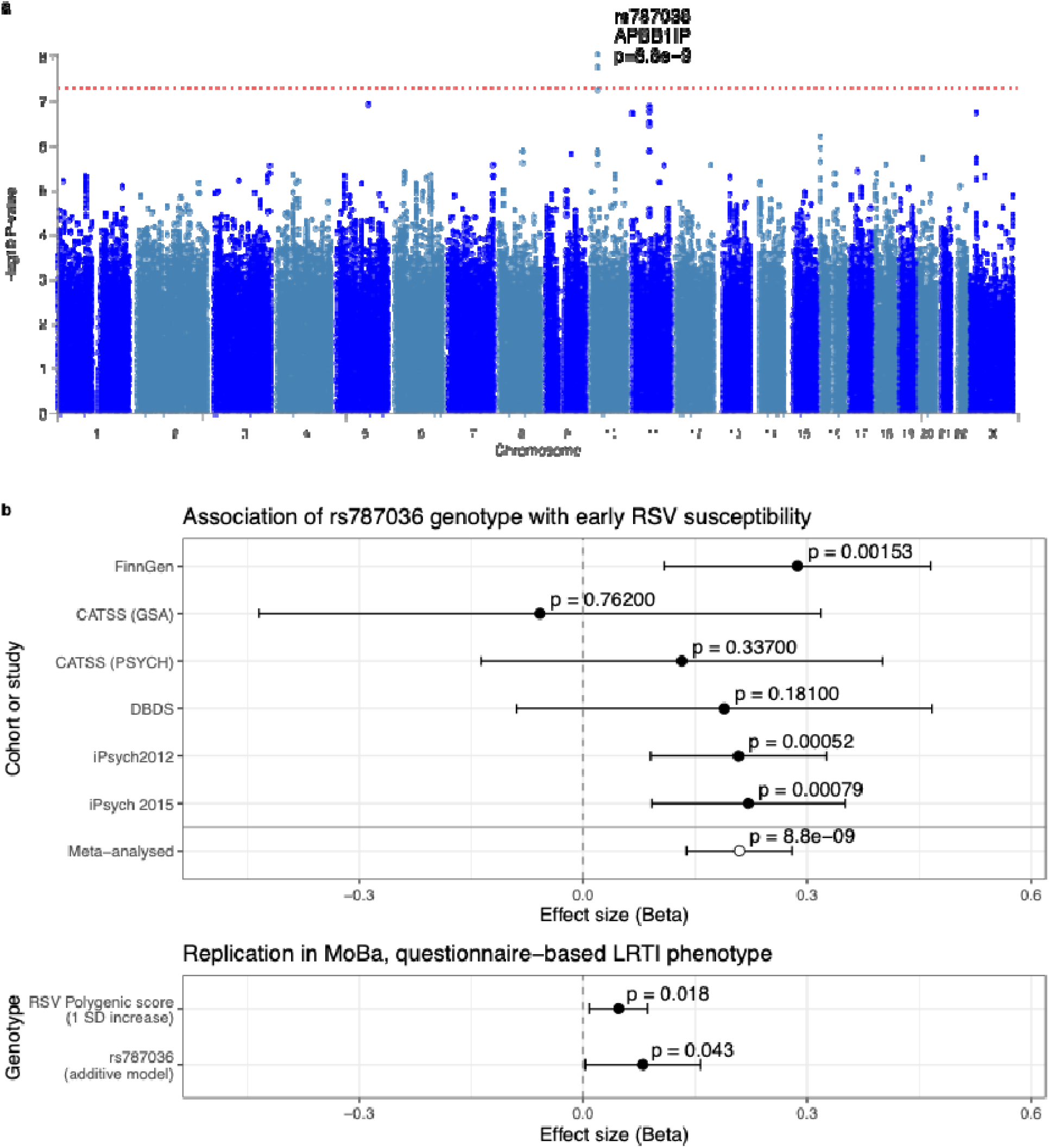
**Panel a:** Manhattan plot showing genome-wide significant associations on chromosome 10. The lead variant, with the lowest p value, is annotated. **Panel b:** Association of the lead variant rs787036 with RSV-related phenotypes across discovery cohorts, the meta-analysis, and the external replication cohort. The x-axis shows the beta coefficient from additive genetic models for rs787036 and RSV diagnosis in each cohort and in the meta-analysis. Note that rs787036 was not genotyped in one of the meta-analysed cohorts (Pasanen et al.)^8^, and this cohort is therefore not included here. In the replication cohort MoBa, a diagnosis-based RSV phenotype was not available; therefore, replication was assessed using a questionnaire-based outcome defined as parental report of any LRTI (RSV, bronchitis or pneumonia) by 6 months of age. The RSV polygenic score was constructed from the GWAS meta-analysis summary statistics using the continuous shrinkage (PRS-CS) method. GWAS meta-analysis was done with 3 107 RSV cases and 92 091 controls. The MoBa replication is analysed within 63 341 children with non-missing genotype information and non-missing phenotype, among whom 2 629 were LRTI cases. RSV = Respiratory syncytial virus CATSS = The Child and Adolescent Twin Study in Sweden PSYCH = Illumina PsychChip GSA = Illumina Global Screening Array DBDS = Danish Blood Donor Study iPsych = Integrative Psychiatric Research cohort^10^ SD = Standard deviation

In the sensitivity analysis within FinnGen, there were 467 cases and 22 586 controls in the main inpatient/outpatient RSV phenotype, and 403 cases and 22 650 controls in the inpatient-only RSV phenotype (**Supplementary Figure 6**). These two phenotypes showed near-complete genome-wide concordance (rg = 1·05, SE = 0·07). The lead meta-analysis variant rs787036 showed the same direction and similar effect size in the combined phenotype (beta = 0·287, p = 0·0016) and the inpatient-only phenotype (beta = 0·259, p = 0·013).

### Asthma genetic liability does not causally increase RSV susceptibility

To investigate whether asthma-associated variants influence RSV susceptibility, we selected 155 LD-independent (r²<0·001) genome-wide significant asthma SNPs from a previously published GWAS^7^ and performed a two-sample Mendelian randomization (MR) analysis. The MR analyses provided no statistically significant evidence of a causal effect of asthma liability on RSV infection (inverse-variance–weighted OR 1·04, 95% CI: 0·91–1·19; p = 0·58). Sensitivity analyses consistently yielded null estimates, with no indication of directional pleiotropy (p = 0·64), low heterogeneity (Q(p) = 0·39), and strong instruments (mean F = 74; ***Supplementary Figure 7***). Because the RSV-GWAS identified only weak associations, we were unable to construct a robust genetic instrument to assess the reverse causal direction (RSV liability → asthma). As an additional sensitivity analysis, we repeated the two-sample MR using 84 childhood-specific asthma SNPs^7^ **(Supplementary Table 6)**, which again showed no evidence that genetic childhood asthma liability causally increases RSV susceptibility (**Supplementary Table 7**).

## Discussion

In this study, which integrated population-scale registry phenotyping, within-family comparisons, and genome-wide analyses, children with RSV hospitalisation and subsequent recurrent wheezing/asthma (RW/A) showed a distinct clinical profile, characterised by higher bronchodilator and inhaled corticosteroid use in early childhood but earlier discontinuation after 5 years of age. In addition, these children had lower eosinophil counts than asthmatic children without RSV hospitalisation. In discordant full-sibling pairs, RSV hospitalisation was linked to substantially higher odds of later childhood recurrent wheezing/asthma, while the unaffected siblings also showed increased RW/A risk, consistent with both shared familial predisposing factors and a possible contributory role of RSV itself. The GWAS meta-analysis identified variants in the *APBB1IP* region associated with RSV susceptibility, whereas genetic liability to asthma showed no evidence of a causal effect on RSV risk.

### RSV-associated recurrent wheezing/asthma seems to have earlier onset and earlier discontinuation of inhaled medication

Our population-based comparisons showed greater inhaled medication use in early childhood and reduced use thereafter, suggesting that respiratory morbidity after infant RSV hospitalisation is most prominent in the first years of life. In addition, the lower eosinophil counts observed in children with RSV-RW/A, compared to non-RSV-RW/A, suggest that RSV-associated wheezing symptoms may be less connected to allergic sensitisation. Similar conclusions were reached in data-driven clustering across three cohorts, where early-life RSV bronchiolitis aligned with a “resolving wheeze” endotype,^6^ characterised by a higher early symptom and exacerbation burden but greater resolution over time, in contrast to multiple-trigger asthma in which exacerbations increase over the first three years and then persist. The INSPIRE birth cohort, which prospectively ascertained RSV infection during infancy, similarly reported an age-dependent association between infant RSV infection and childhood asthma, with evidence that the association was stronger for recurrent early-childhood wheeze and non-atopic asthma than for atopic asthma.^3^ Altogether, these findings suggest that infant RSV is more closely linked to transient early wheeze or a less type 2–dominant asthma phenotype than to persistent eosinophilic asthma. Although inhaled corticosteroid purchases were common in early childhood among children with RSV-RW/A, our registry data cannot assess individual treatment indications, response, or prescribing appropriateness. Further studies with more detailed characterisation of wheezing and asthma subtypes and trajectories are needed.

### Within-family analyses support shared familial susceptibility and a possible contribution of severe RSV

In RSV-discordant sibling pairs, the RSV-hospitalised sibling had nearly threefold higher odds of RW/A than the non-hospitalised sibling, and this pattern persisted across strata of parental asthma. The elevated RW/A prevalence seen among the non-hospitalised siblings—twice the population average—indicates increased baseline familial susceptibility to RW/A in families affected by severe infant RSV, arising from inherited factors, shared environment or both. This is also consistent with family history of asthma increasing RSV hospitalisation risk.^19^ Together, these findings support a mixed model: shared familial background, including inherited predisposition and shared environmental factors, may increase the likelihood of both severe RSV and asthma, while severe RSV may also contribute to subsequent asthma or wheezing. Mechanistic studies provide plausibility for both airway injury and longer-lasting immune skewing after RSV,^2^ yet our MR analyses did not support asthma common-variant liability as a driver of RSV susceptibility, suggesting that the shared susceptibility to RSV and recurrent wheezing/asthma may reflect environmental factors, asthma endotypes not captured by the asthma GWAS instruments, or other early-life determinants. The similar antibiotic purchase rates within RSV-discordant siblings **(Supplementary Figure 3)** further argue against broadly increased infection susceptibility as the primary explanation.

Overall, our findings are consistent with a contributory role of severe infant RSV in later asthma. Observational evidence suggests that preventing RSV in the first year of life could reduce early-childhood wheeze/asthma risk,^3^ and repeated RSV-associated lower respiratory infections have been linked to dose-dependent reduction in lung function.^26^ However, randomised RSV prophylaxis trials have not yet shown a clear reduction in childhood asthma,^4,5^ potentially reflecting heterogeneous target populations and non-specific outcomes, or suggesting that preventing severe disease alone may not have the same long-term respiratory impact as preventing RSV infection more broadly. Future prevention trials and post-implementation studies will therefore be most informative if they distinguish prevention of RSV infection from prevention of severe RSV disease, and assess wheezing/asthma trajectories by phenotype.

### Genetic predisposition to RSV appears distinct from asthma genetic liability

Mendelian randomisation did not support that asthma genetic liability causally increases RSV susceptibility. Instead, we identified intronic variants (rs787036, rs1775248, rs812007) at *APBB1IP* associated with RSV susceptibility. *APBB1IP* encodes RIAM, a regulator of integrin activation with downstream effects on immune-cell adhesion, trafficking, and signalling,^27^ and shows higher expression in lymphoid tissues and immune cell populations.^28^

Although we did not identify quantitative trait loci (QTL) evidence for these variants in available public resources or analyse further leukocyte response characteristics, altered integrin activation could plausibly influence RSV severity; hypothetical mechanisms including excess infiltration of neutrophils to the lungs or modification of the T-cell-mediated antiviral responses.^29^ As neutrophil responses to RSV differ between adults and neonates,^30^ the relative lack of broad adult phenotype and QTL associations at rs787036 is consistent with an effect that may be age dependent or restricted to particular immune-cell states. Larger, harmonised RSV GWAS will be important to validate this locus and define its mechanism. Such studies should ideally incorporate more granular severity measures, such as duration and modality of respiratory support, and more generally, it would be interesting to test whether genetic effects differ by age, immune response characteristics, or RSV-associated wheezing/asthma trajectory.

### Strengths and limitations

A key strength of this study is the integration of nationwide, family-linked Finnish registries with genome-wide data, enabling complementary designs at scale: population-level phenotyping, nationwide discordant full-sibling comparisons to reduce shared familial confounding, and an RSV GWAS meta-analysis with independent replication. Together, these approaches address the RSV–asthma association from clinical, familial, and genetic perspectives.

Several limitations warrant consideration. First, exposure and outcomes were registry-based. Our RSV definition captures severe infant disease requiring hospitalisation; milder infections are not ascertained, and the reference group therefore includes children with milder RSV, which likely attenuates effect estimates and restricts inference to severe early-life RSV, as opposed to any RSV infection. Second, RSV hospitalisation overlaps clinically with early wheeze and may partly reflect underlying host susceptibility, genetic or environmental, not fully captured by the present variables. Third, although the recurrent wheezing/asthma reimbursement endpoint (RW/A) is clinically anchored, it is strict and does not capture milder wheezing phenotypes nor different endotypes. Furthermore, although RSV bronchiolitis should not be considered as one of the obstructive episodes required for the present RW/A endpoint, we cannot exclude the possibility of early RSV directly influencing downstream asthma treatment and reimbursement. Fourth, the sibling design reduces but cannot eliminate residual non-shared confounding. Finally, despite meta-analysis, replication, and consistent direction at the lead locus, the RSV GWAS remains modest in case numbers, and most participants were from the Nordic countries and of European ancestry, limiting generalisability to populations with different genetic ancestries but also potentially to regions with differing RSV epidemiology.

## Conclusions

We combined nationwide registries, within-family comparisons, and genome-wide analyses to clarify the relationship between RSV and asthma. Their association is unlikely to be explained primarily by shared asthma genetic liability, as known asthma-associated genetic variants were not associated with risk of severe RSV, and remains compatible with contributions from both familial susceptibility and RSV. RSV-associated recurrent wheezing/asthma in childhood showed earlier initiation and earlier discontinuation of inhaled therapy and lower blood eosinophils. We identify a genome-wide significant RSV susceptibility locus with supportive evidence in an external replication cohort, motivating larger, harmonised RSV GWAS and mechanistic studies to define causal pathways and susceptible subgroups. These findings help prioritise long-term respiratory outcomes for RSV prevention trials and post-implementation monitoring.

## Contributorship statement

PV, SH, and AG conceptualised and initiated the study. PV was the study coordinator and lead analyst. TH, KD, EV, and MF performed the preprocessing of the Finnish Registry data; PV, with assistance from AV, TH and MP, conducted the Finnish registry-based analyses. MP is the principal investigator for the Finnish registry data used in this study. YL, MCM, and principal investigators SEH and PM provided expertise and data management for the MoBa cohort, while YL and PV ran the replication analyses. TG, GD, SR, CL, and principal investigator CA coordinated the CATSS analyses. CE, SRO, OBP, and ES led the DBDS study and contributed to the GWAS meta-analysis. AEC, TKF, AP, and MR provided summary statistics from existing GWAS publications and contributed to the design of the genetic analyses. HH (FinnGen), MAM (DBDS), TG (CATSS) and YL (MoBa) performed the genetic analyses in respective cohorts; HH conducted the GWAS meta-analysis and PV ran the MR analyses. AJ and AG provided advisory oversight for all genetic analyses, and TL provided clinical expertise related to asthma. PV drafted the manuscript with critical revisions and input from all authors. PV had final responsibility for the decision to submit for publication. All authors have read and approved the final version of the manuscript. PV serves as the guarantor and accepts full responsibility for the work and the conduct of the study. Due to the sensitive nature of the study data, personal data access was only sought and granted for those researchers personally doing data analysis or preprocessing. Principal investigators of each included cohort have verified and are responsible for the underlying data reported in this study.

## Data sharing

Individual-level data of this study cannot be shared. Meta-analysed summary statistics from the RSV GWAS meta-analysis is deposited with the GWAS Catalog (https://www.ebi.ac.uk/gwas/home) with study accession code GCST90860867. Access to Finnish registry data can be obtained by submitting a data permit application for individual-level data to the Finnish social and health data permit authority; Findata (https://www.findata.fi/). Requests are evaluated on a case-by-case basis. The code of this project is available in GitHub (https://github.com/dsgelab/rsv_asthma).

## Supporting information

STROBE-MR

STREGA

## Data Availability

Individual-level data of this study cannot be shared. Meta-analysed summary statistics from the RSV GWAS meta-analysis will be deposited with the GWAS catalog (https://www.ebi.ac.uk/gwas/home). Access to Finnish registry data can be obtained by submitting a data permit application for individual-level data to the Finnish social and health data permit authority; Findata (https:/www.findata.fi/). Requests are evaluated on a case-by-case basis.

## Acknowledgements

We are grateful to children and their families whose data made this study possible. This study and PV were supported by the Päivikki and Sakari Sohlberg Foundation, the Foundation for Pediatric Research, the Orion Research Foundation and the Sigrid Jusélius Foundation (grant number 220024). We acknowledge CSC - IT Center for Science, Finland for computational resources. PV acknowledges the use of large language models (OpenAI ChatGPT and Google Gemini) for language editing. AG acknowledges the research environment provided by ELLIS Institute Finland. We also acknowledge the Swedish Twin Registry for access to data. The Swedish Twin Registry is managed by Karolinska Institutet in Stockholm, Sweden, and is partially funded by the Swedish Research Council under the grant numbers 2017-00641 and 2021-00180. We also thank the Biobank at Karolinska Institutet for professional biobank service. Financial support was also provided by the Swedish Research Council (grant no 2023-02327), the Swedish Heart-Lung Foundation (grant no 20240974), the Swedish Asthma and Allergy Association Research Fund (grant no 2024-0010), and grants provided by Region Stockholm (ALF project RS2022-0674). The study was also partly supported by the Research Council of Norway through its Centres of Excellence funding scheme (project No 262700). The DBDS wishes to thank the Danish blood donors for their contribution and support, and Amgen deCODE Genetics for genotyping the cohort. The initiation of DBDS was supported by the Danish Administrative Regions (02/2611) and the Danish Council for Independent Research (09–069412). Additionally, DBDS is partly funded by Bio- and Genome Bank Denmark, the Novo Nordisk Foundation (NNF23OC0082015, NNF17OC0027864, and NNF17OC0027594) and EU Horizon (grant no 101057129).

