## Supplementary figure for "Infant respiratory syncytial virus and childhood asthma: a nationwide registry-based, sibling-controlled, and genome-wide association study"

Supplementary material

### Registry-based cohorts and data

##### Finnish registry cohorts and family linkage (FinRekisterit2025 project)

We used data from nationwide health and administrative registries in Finland. The Finnish registry-based analyses were based on the FinRekisterit2025 project, coordinated by the Finnish Institute for Health and Welfare (THL). The FinRekisterit2025 was approved by THL (decisions THL/5757/6.02.00/2025; THL/2844/6.02.00/2023 and THL/6366/6.02.00/2024).

FinRekisterit2025 is a nationwide, registry-based data resource covering all individuals living in Finland on 1 January 2010 (index persons), as well as their parents, spouses, children and siblings, altogether approximately 7.2 million individuals. See in-depth description of data, original registries and preprocessing steps in the cohort profile.^1^ Individual-level data from multiple national registries are linked using the unique (pseudonymised) personal identity code.

Familial relationships were obtained from the Digital and Population Data Services Agency (DVV), enabling construction of multigenerational family links. For the present study, FinRekisterit2025 provides the sampling frame and follow-up structure for identifying RSV-related hospitalisations, relevant covariates and subsequent asthma outcomes in the Finnish population.

##### Finnish Medical Birth Registry

Detailed perinatal and maternal data was obtained from the Finnish Medical Birth Registry, which has collected information since 1987 on all births in Finland, including maternal characteristics, pregnancy and delivery details, and neonatal diagnoses and outcomes. In this study, Hilmo and the Medical Birth Registry were used to define early-life RSV hospitalisations, birth characteristics and perinatal covariates.

##### Hospital care data

Information on hospital-based care is derived from the Finnish Care Registry for Health Care (Hilmo), which includes nationwide data, including all diagnoses, on inpatient hospitalisations and specialised outpatient visits. The data is considered to be nationwide and of high quality,^2^ especially on paediatric hospitalisations. Presently, only International Classification of Diseases, 10th revision (ICD-10) codes were used, due to all study subjects being born after the implementation of ICD-10 in Finland.

##### Medication data (Kela drug purchases and reimbursements)

Medication use is captured through two complementary national registries. First, the reimbursement and purchase of prescription medicines are recorded in the nationwide drug purchase registries maintained by the Social Insurance Institution of Finland (Kela), which have collected reimbursable outpatient prescriptions since 1995 and special reimbursement entitlements for chronic diseases (including asthma) since 1968. These registries include Anatomical Therapeutic Chemical (ATC) codes, package information and costs, and are used to define long-term medication use and disease-specific reimbursement status.

##### Kanta Laboratory data

In Finland, laboratory test results are stored as part of the Kanta Services Patient Data Repository, the national electronic health record archive that aggregates clinical records from both public and private healthcare providers. Laboratory results recorded to Kanta are visible to citizens via MyKanta, and for secondary use in research can be accessed via the Finnish data permit authority Findata under the Secondary Use framework. Kanta data collection started in 2014, with healthcare providers gradually joining the system.

A practical consideration for research is that Kanta laboratory data is heterogeneous: results are stored under the laboratory view using multiple coding systems (including local laboratory code systems), meaning that the same test may appear under different codes and names across providers and time; in a sample-based overview, hundreds of distinct laboratory code systems were observed. Presently, we identified eosinophil values coded with the most common laboratory codes and names used for absolute eosinophil counts, and which were recorded in cells *10^9 / Litre.

Kanta laboratory data was not fully nationwide throughout the study period, and therefore do not capture all eosinophil measurements for all children. In the earlier years of follow-up, inclusion of a laboratory result in Kanta depended on the healthcare provider’s timing of adoption of the Kanta Patient Data Repository, leading to incomplete and potentially geographically varying coverage. In practice, larger public providers (including paediatric hospitals) contributed laboratory results earlier, which may introduce selection related to region and care sector (public vs private) among children with available measurements. However, our analyses were restricted to eosinophil values recorded within Kanta, and eosinophil comparisons were made only between children whose data was in Kanta, reducing sensitivity to differences in coverage across providers. For an overview of Kanta data resources (including also other modalities than laboratory measurements), see the preprint by Männikkö et al.^3^

#### Missing or incomplete data in Finnish registries

Missingness in routinely collected registry data in Finland is very rare. Events are coded as having an event of interest, and absence of an event code is by default interpreted as no event. Individual registry variables in the Finnish Medical Birth Registry contain some missing values.

Among the full-sibling sub-cohort of 527 776 subjects, the following variables had missing values, and cases with missingness were excluded from the conditional logistic regression model in which these variables were used as covariates.

Gestational age: 825 (0.16%) of cases
Mother’s smoking during pregnancy: 12 691 (2.4%) of cases

Although very few missing values were identified in the registry data, some of the data used in other analyses may not be fully representative due to incomplete recording.

Notably, among the main study population of 965 312 subjects, 19 814 (2.1%) did not have a recorded father ID variable, and they could not be included in constructing the full-sibling pairs, and whether they had full siblings or not is unknown. These subjects were still included in the population-based analyses.

As described above in the Kanta Laboratory section, eosinophil data does not cover all measurements done in Finland, but as we only restricted to comparing cases with existing data, we estimate that non-representativeness does not have that large impact on within-group differences, but the absolute eosinophil values and magnitude of differences might not be representative of the full populations.

Also in the inhaled medication purchase and eosinophil trajectories, we extended the follow-up to 12 years of age for descriptive purposes. Some of the children (born between 2009 and 2014) did not have complete registry-based follow-up during the ages 7 to 12. We deemed this acceptable since the main aim of visualising the trajectories was descriptive, and the majority of children (those born between 1998 and 2009) had complete follow-up also until the age of 12 years.

With regard to genetic analyses, missingness was not considered and only subjects with existing genotype and outcome data were included in the analyses, as is the standard practice.

### Genotyped cohorts

#### FinnGen

FinnGen is a biobank collaboration project which includes subjects and samples from hospital biobanks and prospective epidemiological and disease-based cohorts^4^ (https://www.finngen.fi/en). Approximately 10% of the Finnish population are participants in FinnGen. Samples were genotyped with Illumina (Illumina) and Affymetrix arrays (Thermo Fisher Scientific), and imputation is based on a Finnish-specific reference panel; see more details from the original publication.^4^ The data has been interconnected with national registries including hospital discharge records (accessible from 1968), death records (from 1969), cancer registries (from 1953) and drug purchase records (from 1995), primary and secondary care diagnoses, and many disease-specific registries. FinnGen phenotype data has been curated to clinically relevant disease endpoints, which combine e.g. several diagnoses (including harmonisation across different ICD systems), drug purchases and operation codes into clinically relevant disease definitions. FinnGen Data Release 12 was used in this study, which includes 520 210 genotyped individuals. The RSV-GWAS analysis was specifically approved by the FinnGen scientific committee (Proposal number F_2024_065, approved in 10.7.2024).

#### Danish Blood Donor Study (DBDS)

The Danish Blood Donor Study (DBDS)^5^ was established in 2010 to study the impact of blood donation on the health of blood donors and to answer broader health-related research questions. By 2015 it was nationwide. The genotyping was done with Infinium Global Screening Array (Illumina Inc., USA), performed at deCODE genetics (Reykjavik, Iceland). Performing genetic analyses in DBDS was approved by the National and Zealand Committees on Health Research Ethics, NVK-1700407 and SJ-740, and Capital Region Data Authority, P-2019-99. Currently the cohort comprises more than 175 000 individuals, of which 135 000 have been genotyped. Participants’ data is linked to national health registries, providing e.g. prospective and longitudinal data on hospital contacts, diagnoses, prescriptions and more. In addition, DBDS contains a wealth of biomarker and laboratory data, and also self-reported data.

#### The Child and Adolescent Twin Study in Sweden (CATSS)

Child and Adolescent Twin Study in Sweden **(CATSS)** is an ongoing, population-based longitudinal cohort within the Swedish Twin Registry that targets all twins born in Sweden since July 1, 1992.^6^ Notably, the cohort consists of twins. Relevant for the present study, data has been linked to national diagnosis registry records. CATSS participants were genotyped on two SNP arrays (Illumina PsychChip and Illumina Global Screening Array, GSA).

#### Previously published genome-wide association studies of RSV contributing summary statistics for the meta-analysis

To our meta-analytic RSV GWAS, we included summary statistics from two existing publications of GWAS on RSV susceptibility.

**iPSYCH**
The Integrative Psychiatric Research (iPSYCH) study is a large, population-based Danish case–cohort sample established to investigate the genetic and environmental architecture of severe mental disorders. It consists of two sub-cohorts, iPSYCH 2012 and iPSYCH 2015, both including subjects with psychiatric diagnoses and randomly selected controls from the general population. A publication in iPSYCH cohort^7^ did a GWAS on RSV susceptibility, using RSV hospitalisation in ages 0 to 5 years as the outcome, was the largest previously published GWAS with 1786 cases and 45 060 controls.

For both iPSYCH2012 and iPSYCH2015, DNA was extracted and whole-genome genotyping performed from neonatal dried blood spots stored in the Danish Neonatal Screening Biobank. iPSYCH2012 samples were genotyped using the Illumina PsychChip, with successful genotyping in approximately 90% of individuals, while the iPSYCH2015 expansion was genotyped using the Illumina Global Screening Array. All participants are linkable, via the unique personal identifier assigned to Danish residents, to nationwide health and administrative registries. The Danish National Patient Register (DNPR) was used in the original study to extract information on RSV hospitalisations, coded with ICD-10 codes after the year 1995. The RSV-GWAS was done separately in the two cohorts, iPSYCH2012 and iPSYCH2015. We obtained summary statistics from both of these analyses separately and combined them in our present larger meta-analysis. See original iPSYCH cohort descriptions for more details.^8,9^

**The publication by Pasanen et al.**

We included summary statistics from a genome-wide association study of viral bronchiolitis conducted in Finnish, Swedish, and Dutch infants.^10^ The discovery GWAS (included in the present GWAS meta-analysis) comprised 217 bronchiolitis cases and 778 controls from Finland and Sweden. Cases were required to have clinician-diagnosed bronchiolitis before 24 months of age, defined as lower respiratory tract infection with typical bronchiolitis symptoms (wheezing and laboured breathing). Controls were population-based children without a history of bronchiolitis, frequency-matched by age and region. Genomic DNA was extracted from whole blood, buffy coat, or buccal swabs, and genome-wide genotyping was performed at the Technology Centre of the Institute for Molecular Medicine Finland (FIMM) using Illumina Human Genotyping Array Kits, followed by standard quality control and imputation with SHAPEIT2 and IMPUTE2 to a dense reference panel (>5.3 million SNPs with MAF >0.01 available for analysis). See original publication for details.^10^

#### Norwegian Mother, Father and Child Cohort study (MoBa) as the replication cohort

The Norwegian Mother, Father and Child Cohort Study (MoBa) is a population-based pregnancy cohort established by the Norwegian Institute of Public Health, with recruitment through routine antenatal care in Norway from 1999–2008 and a participation rate of approximately 41%. The cohort comprises ~114 000 children, ~95 000 mothers and ~75 000 fathers, and includes a family design with biological samples collected from both parents during pregnancy and from mothers and children (from cord blood sample) at birth. Questionnaires have been administered to parents and later to children throughout the participants’ life. For more details, see the most recent cohort description.^11^ Genotype data in MoBa have been generated across several large genotyping efforts, using DNA from the biobank and Illumina microarray platforms, including HumanCoreExome, Global Screening Array (GSA), and OmniExpress/Infinium OmniExpress arrays (among others), with downstream quality control and imputation in the MoBaGenetics resource. These analyses were carried out as part of the project “Den maternelle effekten på barneastma”, project ID 2495483.

In this study, MoBa was used as an independent replication cohort for lead variants from the RSV susceptibility GWAS meta-analysis; because linked clinical diagnosis data was not available for the replication phenotype, RSV susceptibility was defined using the parental report on any lower respiratory tract infection (RSV, bronchitis or pneumonia) during the first 6 months of life.

In MoBa, the number of genotyped participants with non-missing LRTI outcome data was 63 341. Among 111 921 pregnancies in which mothers originally enrolled, 87 917 returned valid responses to the LRTI-related questions at 6 months. For 63 537, we could calculate a valid polygenic score for RSV, and finally the number of persons with non-missing genotype in rs787036 and non-missing polygenic score (PGS) was 63 341.

### Genetic analyses

#### RSV Phenotype definition for GWAS meta-analysis

We provided the following definition to participating studies for them to define RSV cases and controls.

##### Case definition

All of the following criteria 1-3 should be met

1. Any of the following diagnoses (as either inpatient or outpatient)
   a. **ICD-10**:
      i. J21.0 (Acute bronchiolitis due to respiratory syncytial virus)
      ii. J12.1 (Respiratory syncytial virus pneumonia)
      iii. J20.5 (Acute bronchitis due to respiratory syncytial virus)
   b. **ICD-9**: 466.10, 480.1 (note that all participating cohorts had ICD-10 coding system for the birth years included)
2. Age <2 years at time of event
3. Born after 01-01-1995

##### Control definition

1. Born after 01-01-1995

Studies will be considered eligible for this meta-analysis study if the number of cases in RSV phenotype was >= 50.

The birth year definition (after 1995) for both cases and controls was set to ensure that each participant has been born during time when RSV testing had been implemented to paediatric emergency care units, which in turn would increase the specificity of the listed RSV diagnoses.

##### Phenotype differences in previously published studies

In the two published studies included in the GWAS meta-analysis, the phenotype was defined only to include RSV hospitalisations. In the iPSYCH cohort, the phenotype was RSV hospitalisation with ICD-10 diagnoses above, at or before the age 5 years. In the publication by Pasanen et al., the phenotype was RSV diagnosis (both in- and outpatients) at or before the age of 2 years.

We accepted this difference, since most medically attended RSV infections occur during the first year of life, and cases occurring after 2 years are infrequent.^7^ Related to the inpatient-outpatient setting, we accepted this difference of some cohorts not contributing outpatient (non-hospitalised) RSV cases, since disease severity in RSV is a continuum and only partly and imperfectly captured by the binary inpatient/outpatient classification.

##### Sensitivity analysis of inpatient-only RSV phenotype in FinnGen

In the FinnGen cohort, we did a sensitivity analysis by running a GWAS using an inpatient-only RSV case definition and compared it with the primary FinnGen GWAS (including both in- and outpatient RSV cases. We assessed the concordance of genetic signal (ie, genetic correlation) between the two phenotypes with Linkage Disequilibrium Score Regression (LDSC) and by comparing effect estimates for the genome-wide significant variants from the main meta-analysis.

#### Statistical genetic analysis methods and quality control

##### Models used to test for genetic association

In the GWAS analysis, all participating cohorts used the following standard association model:

Outcome ~ variant + birth year + sex + birth year*sex + PCs + [study-specific covariates]

We suggested not to exclude related individuals but instead correctly model relatedness using mixed model-based software packages (e.g. BOLT-LMM, SAIGE, REGENIE).

All participating cohorts consisted mainly of persons with European ancestry, but we instructed to run the GWAS separately for each major ancestry group (only European-ancestry summary statistics were returned). We recommended adjusting for 20 Principal Components (PC). *Study_specific_covariates* indicates covariates used to correct technical artifacts (e.g. batch number) and not risk factors or other comorbidities.

##### Genotypes & imputation in individual studies

We recommended using imputed genotypes for analyses. For genotype imputation, participating studies used their local established practices, and for details for imputation we kindly refer to the original cohort descriptions. For consistency, in the analysis plan, we recommended using either a local specific reference panel, existing imputation panels or use the TopMed imputation server (<https://imputation.biodatacatalyst.nhlbi.nih.gov/>) or the Michigan Imputation Server (<https://imputationserver.sph.umich.edu/index.html>).

For processing of genotyping data, we recommended using the Rapid Imputation for COnsortias PIpeLIne (RICOPILI)^12^ pipeline (<https://sites.google.com/a/broadinstitute.org/ricopili/>). Recommendations for quality control (QC) parameter specifics can be found at <https://sites.google.com/a/broadinstitute.org/ricopili/preimputation-qc#TOC-Technical-Details>, and are reported below for simplicity. We allowed minor adjustments for the parameters according to the local characteristics of the data if appropriate, but no deviations were reported.

- Sample QC: call rate in cases or controls ≥ 0.98
- Sample QC: FHET within +/- 0.20 in cases or controls
- Sample QC: Sex violations (excluded) - genetic sex does not match pedigree sex
- SNP QC: call rate ≥ 0.95
- SNP QC: missing difference ≤ 0.02
- SNP QC: Hardy-Weinberg equilibrium (HWE) in controls p value ≥ 1e-06 (i.e., ≥ log 10(p) of -6)
- SNP QC: Hardy-Weinberg equilibrium (HWE) in cases p value ≥ 1e-10 (i.e., ≥ log 10(p) of -10)
- (HWE step should be done only in females when applying to chromosome X)

Before the meta-analysis, the summary statistics were standardised, filtered (excluding variants with allele frequency <1% or imputation INFO score <0.6), lifted over to reference genome build GRCh38 (in studies imputed to GRCh37).

Meta-analysis was carried out in human genome reference build hg38, submissions accepted either in hg19 or hg38. All results are given in hg38.

#### Meta-analysis of GWAS results

After receiving the GWAS summary statistics, we did a meta-analysis with summary statistics from the participating studies (FinnGen, CATSS, DBDS) and the two earlier publications.

Meta-analysis was done with the METAL software^13^ (<https://csg.sph.umich.edu/abecasis/Metal/>, version 2011/03), using the inverse-variance fixed-effect (or standard error) scheme. We also tested for heterogeneity of variant effects estimates across studies with Cochran’s Q-test and I2 (the percentage of variation due to actual heterogeneity rather than chance). Cutoff for genome-wide significance was 5e-8.

#### Polygenic risk score for RSV

In order to test whether genetic susceptibility to RSV replicates in other cohorts also beyond the genome-wide significant SNPs, we constructed a polygenic score (PGS) for RSV based on the GWAS summary statistics. Here we used the PRS-CS method.^14^ It applies Bayesian continuous shrinkage (CS) priors to improve the accuracy of PGS, particularly in cases of small sample sizes or when dealing with genetic variants of small effect sizes. PRS-CS uses a normal prior for SNP effects and automatically estimates the optimal shrinkage parameters, which enhances the model's predictive power. This method is particularly effective for complex traits, as it incorporates both genome-wide data and the continuous nature of effect sizes across the genome.

#### Downstream analysis of genetic associations

To assess possible genetic correlations of RSV and other phenotypes, we used the Linkage Disequilibrium score regression method.^15^ We searched for associations of the lead SNP (rs787036) from genetic association databases: <https://platform.opentargets.org/>, [results.finngen.fi](http://results.finngen.fi), <https://pheweb.jp/>, <https://pheweb.org/UKB-Neale/>. We also used a tool [anno.finngen.fi](http://anno.finngen.fi) (requires FinnGen account), that allows searching for association results (including quantitative trait loci mappings, QTLs) from multiple biobanks’ summary statistics. We also browsed the Genotype-Tissue Expression (GTEx) project resources, including the GTEx Portal (<https://gtexportal.org/home/>), and checked for methylation QTLs from <http://www.mqtldb.org/>. We also explored the variant effects on splicing by using <https://spliceailookup.broadinstitute.org/> database.^16^

##### Two-sample mendelian randomisation

The main GWAS-based analysis was two-sample mendelian randomisation (MR). With this we aimed to investigate the shared genetic architecture of RSV and asthma.

We selected approximately independent asthma loci using linkage disequilibrium (LD) clumping. Starting from the largest asthma GWAS summary statistics,^17^ we retained variants with genome-wide significance (P < 5×10⁻⁸) and valid rsIDs. Independence was defined by PLINK (v.1.9.0) clumping using a European-ancestry LD reference panel. We used a 10 Mb window and an LD threshold of r² < 0.001, with the index variant threshold set at P < 5×10⁻⁸. For each locus, the most significant SNP was kept as the lead (‘index’) variant and all other variants within ±10 Mb in LD above the threshold were removed. Of these 160 SNPs, we excluded 1 SNP due to incompatible alleles and 4 due to palindromic SNP labelling and intermediate allele frequencies in comparison with the RSV-GWAS summary statistics. The resulting set of 155 independent asthma SNPs was then tested for association with the RSV phenotype. See list of SNPs in **Supplementary Table 5**.

Causal effects were estimated using inverse-variance weighting as the primary method, with MR-Egger, weighted median and weighted mode as sensitivity analyses; pleiotropy and model validity were assessed using the MR-Egger intercept, Cochran’s Q for heterogeneity, and instrument strength (F-statistics). We used R v. 4.5.0 and package TwoSampleMR v.0.6.22 to carry out two-sample mendelian randomisation (MR).

As an additional sensitivity analysis, we ran the two-sample mendelian randomisation by using only childhood-onset asthma (COA) -specific SNPs. In the original asthma GWAS,^17^ the authors carried out a supplementary analysis of investigating COA-specific effects in comparison to adult-onset asthma, in those cohorts where COA could be defined. For our sensitivity analysis, we selected 84 SNPs that were genome-wide significant after harmonisation in the COA-specific meta-analysis. See the list of childhood-specific SNPs in **Supplementary Table 6**, and COA-specific MR in **Supplementary Table 7.**

The two-sample MR analysis relies on the standard instrumental variable assumptions. First, the relevance assumption requires that the selected variants are robustly associated with the exposure, asthma genetic liability. We addressed this by selecting genome-wide significant, LD-independent asthma-associated variants from the largest published asthma GWAS and by reporting instrument strength using F-statistics. The independence assumption requires that the genetic instruments are not associated with confounders of the relationship between asthma liability and RSV susceptibility. This assumption cannot be fully tested. Potential bias from population structure was partly accounted for by using ancestry-adjusted GWAS summary statistics, as is standard practice; furthermore, both the exposure and outcome GWASs were done on cohorts mostly of European ancestry. However, pleiotropic associations of asthma variants with early-life factors related to RSV risk cannot be excluded, although we think they are very unlikely (most important RSV risk factors being gestational age, young age during infection, having siblings in family, and certain congenital diseases). Third, the exclusion restriction assumption requires that the instruments influence RSV susceptibility only through asthma liability and not through alternative biological pathways. This assumption cannot be proven empirically; therefore, we assessed potential violations using MR-Egger intercept, Cochran’s Q, weighted median and weighted mode sensitivity analyses, and leave-one-out analyses.

We did not use individual-level data from the asthma GWAS and could not quantify sample overlap between the published asthma exposure GWAS and the RSV outcome GWAS. However, the asthma exposure GWAS was very large with >150 000 cases across 22 biobanks, and any possible overlap is expected to be very limited, and the MR was performed using strong external asthma instruments.

### Supplementary Figures

#### S1: Eosinophil trajectory model accounting for repeated measurements

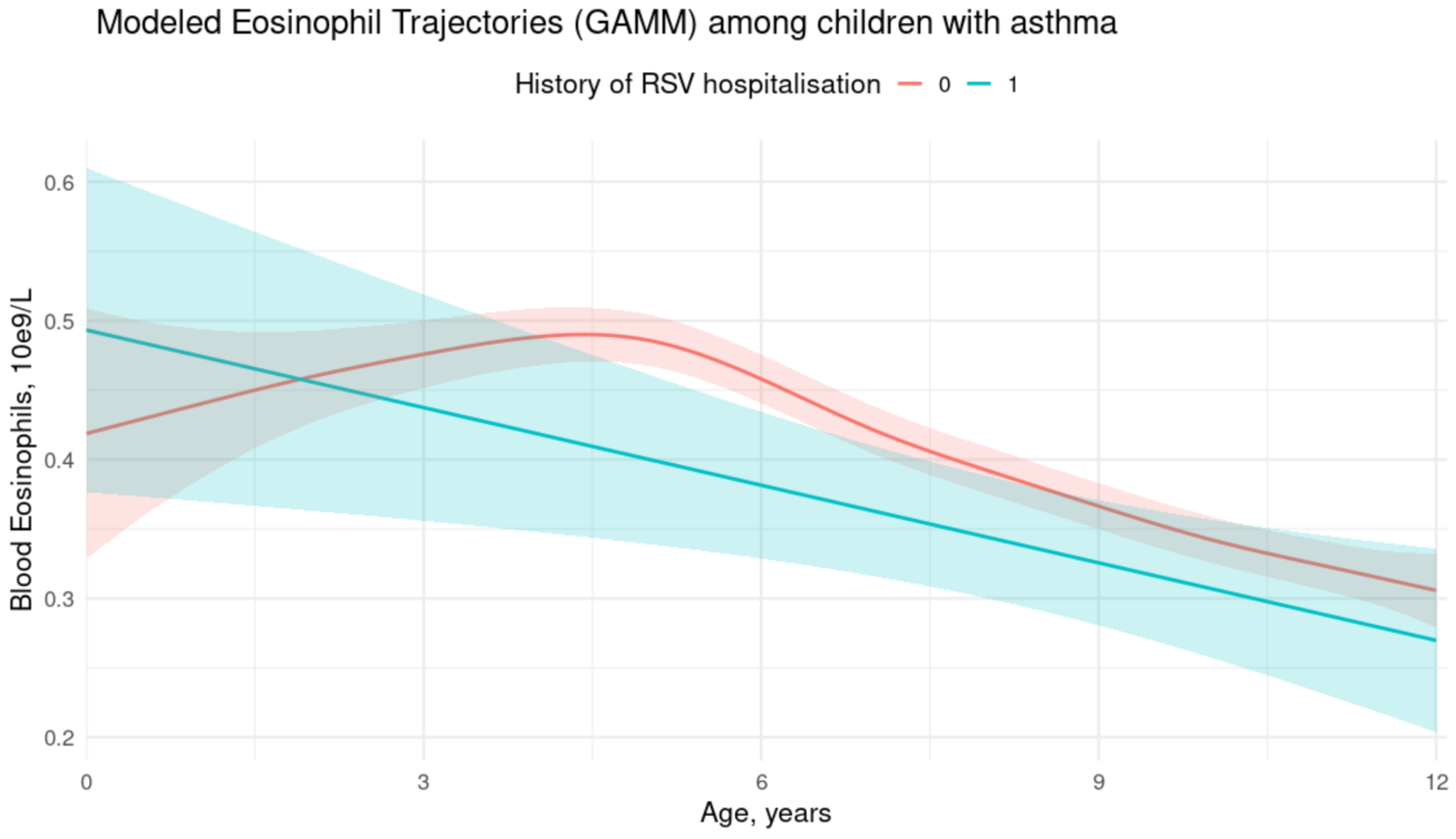

**Supplementary Figure 1**: Eosinophil Trajectories Modeled Using a Generalised Additive Mixed Model (GAMM). The y axis shows blood eosinophil count in 10e9 / L. This analysis, performed only among children with childhood-onset asthma (COA) for computational efficiency, compares eosinophil trajectories between children with recurrent wheezing/asthma (RW/A) and history of RSV hospitalisation (RSV-RW/A) and those with RW/A but no RSV hospitalisation (non-RSV-RW/A). The model accounts for repeated eosinophil measurements from the same child by introducing a random intercept for each individual, ensuring that the analysis captures within-subject correlation of different measurements. The observed trend of higher eosinophil counts in children with COA but no RSV hospitalisation is consistent with the main analysis (**Figure 3**). The GAMM was fitted using the gamm() function from the mgcv R package. Ribbons show 95% confidence intervals for predicted eosinophil values.

For computational constraints (GAMM models are computationally intensive), and due to RSV-COA vs non-RSV-COA being the comparison of interest, this analysis was done only among children with COA, using 6 588 measurements from 4367 children with non-RSV-COA and 402 measurements from 248 children with RSV-COA.

#### S2: Directed acyclig graph (DAG) of confounders in the sibling model

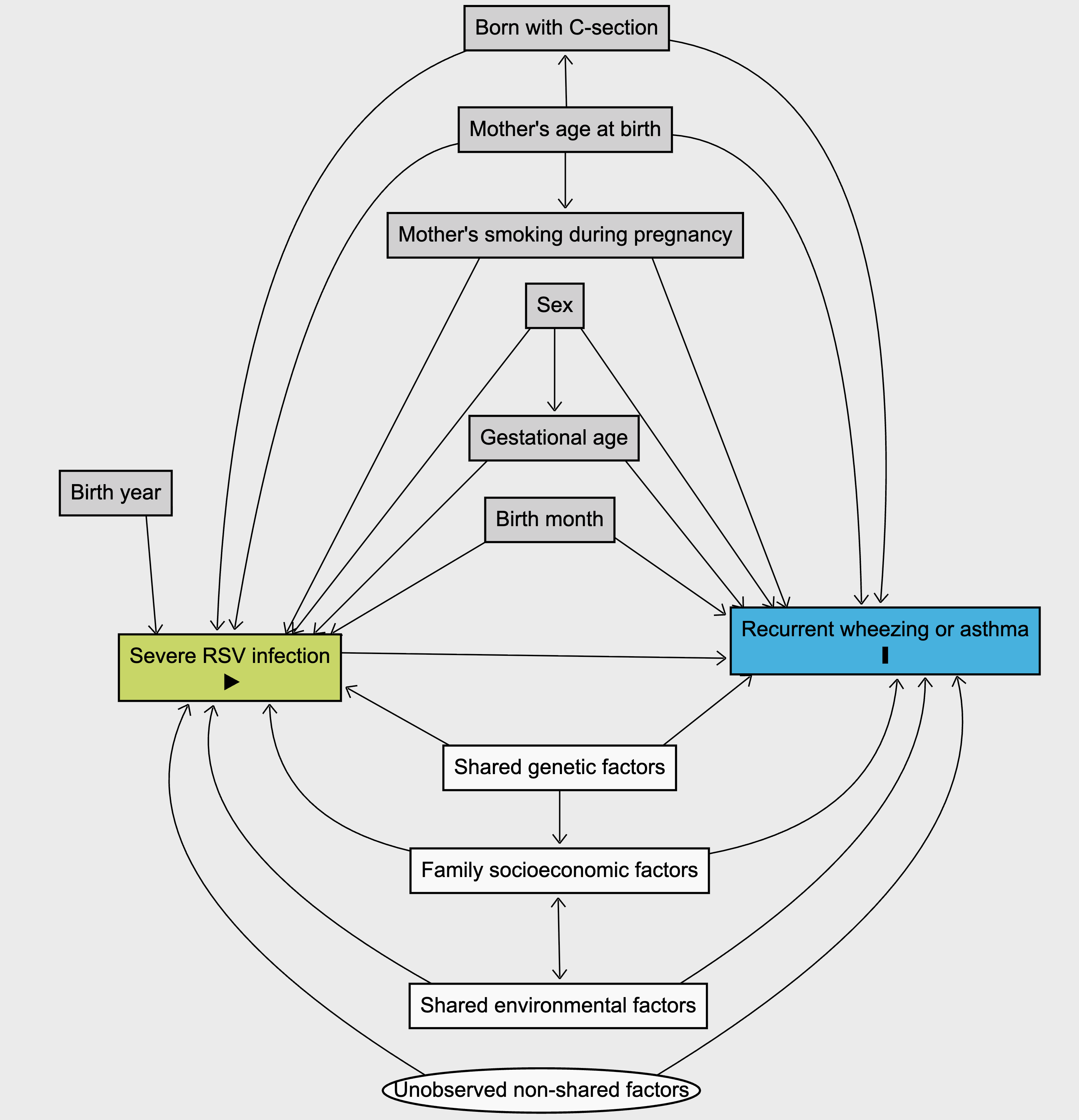

**Supplementary figure 2**: Simplified DAG showing the association and confounders in the conditional logistic model of discordant full sibling pairs. In this model, asthma or recurrent wheezing reimbursement code is the outcome variable (shown in blue). Grey boxes indicate those specific covariates that are potentially not shared between full siblings, that are associated with RSV and/or asthma, and that are included as covariates in the model. White boxes indicate more general factors, that are accounted for by the full sibling analysis setting. Finally, white circle represents residual confounding, i.e. those factors (such as non-shared genetic factors and non-shared environment). that the specific covariates or the full sibling model does not account for.

#### S3: Medication purchases among sibling pairs discordant for RSV hospitalisation

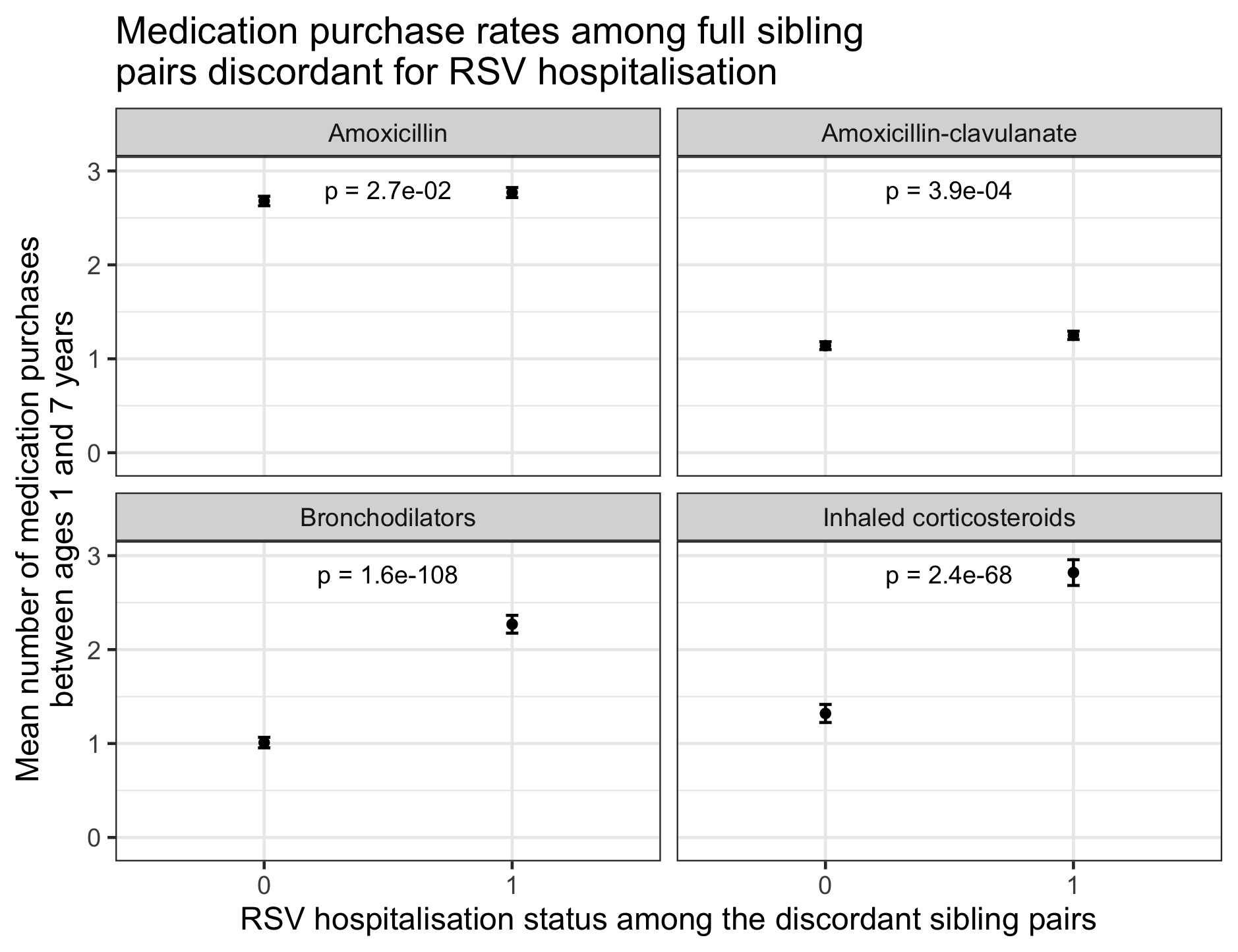

**Supplementary Figure 3:** Medication purchase rates between age 1 and 7 years in RSV-discordant sibling pairs. Panels display purchase rates for two antibiotics (amoxicillin and amoxicillin-clavulanate) and two inhaled medications (bronchodilators and inhaled corticosteroids). Among RSV-discordant siblings, antibiotic purchase rates are very similar, whereas clear differences are observed in bronchodilator and inhaled corticosteroid purchases. P values show the statistical significance of the difference between the two groups (Those with RSV hospitalisation vs their full siblings without RSV hospitalisation during the first year of life). Although the difference in amoxicillin-clavulanate is statistically significant, the absolute difference is very small, in comparison to those observed in inhaled medication purchases.

#### S4: Locus zoom plot on chromosome 10, around the lead SNP region

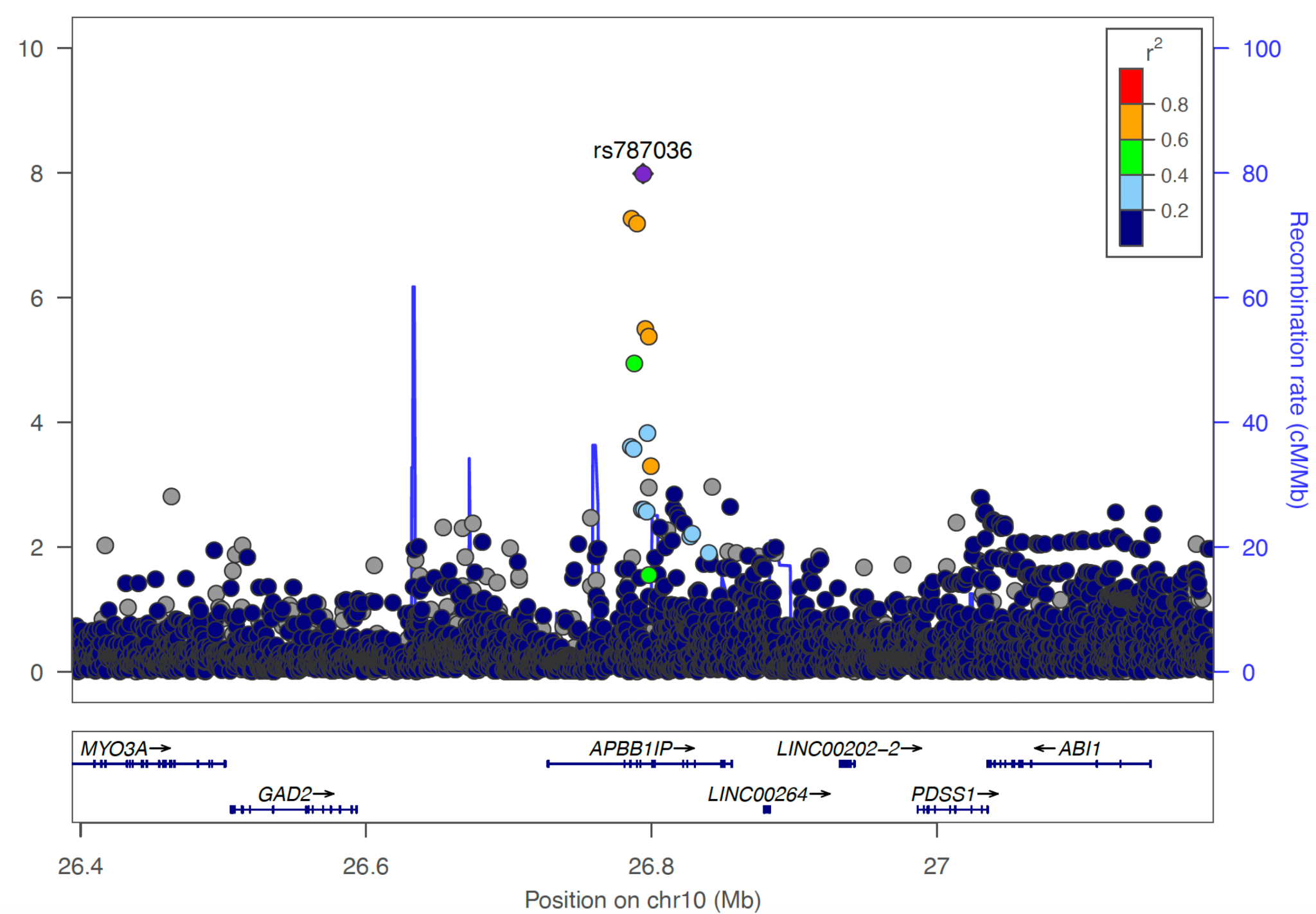

**Supplementary figure 4**: Locus zoom plot of chromosome 10, targeting the region around the lead variant rs787036 in the RSV-GWAS.

#### S5: Q-Q plot

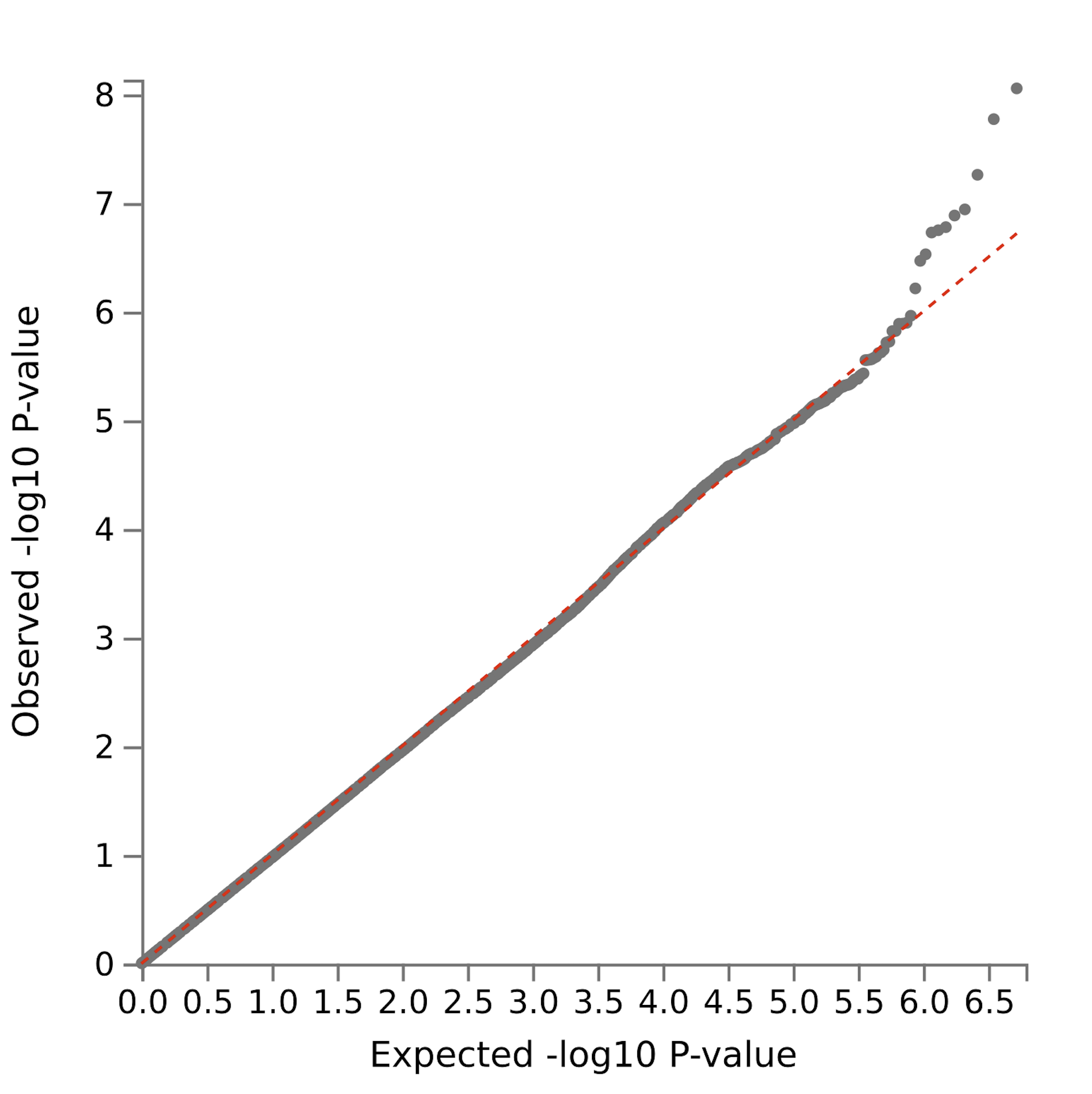

**Supplementary figure 5** The Q-Q plot, showing observed and expected P-values in the RSV-GWAS meta-analysis.

#### S6: Sensitivity analysis of inpatient RSV GWAS in FinnGen

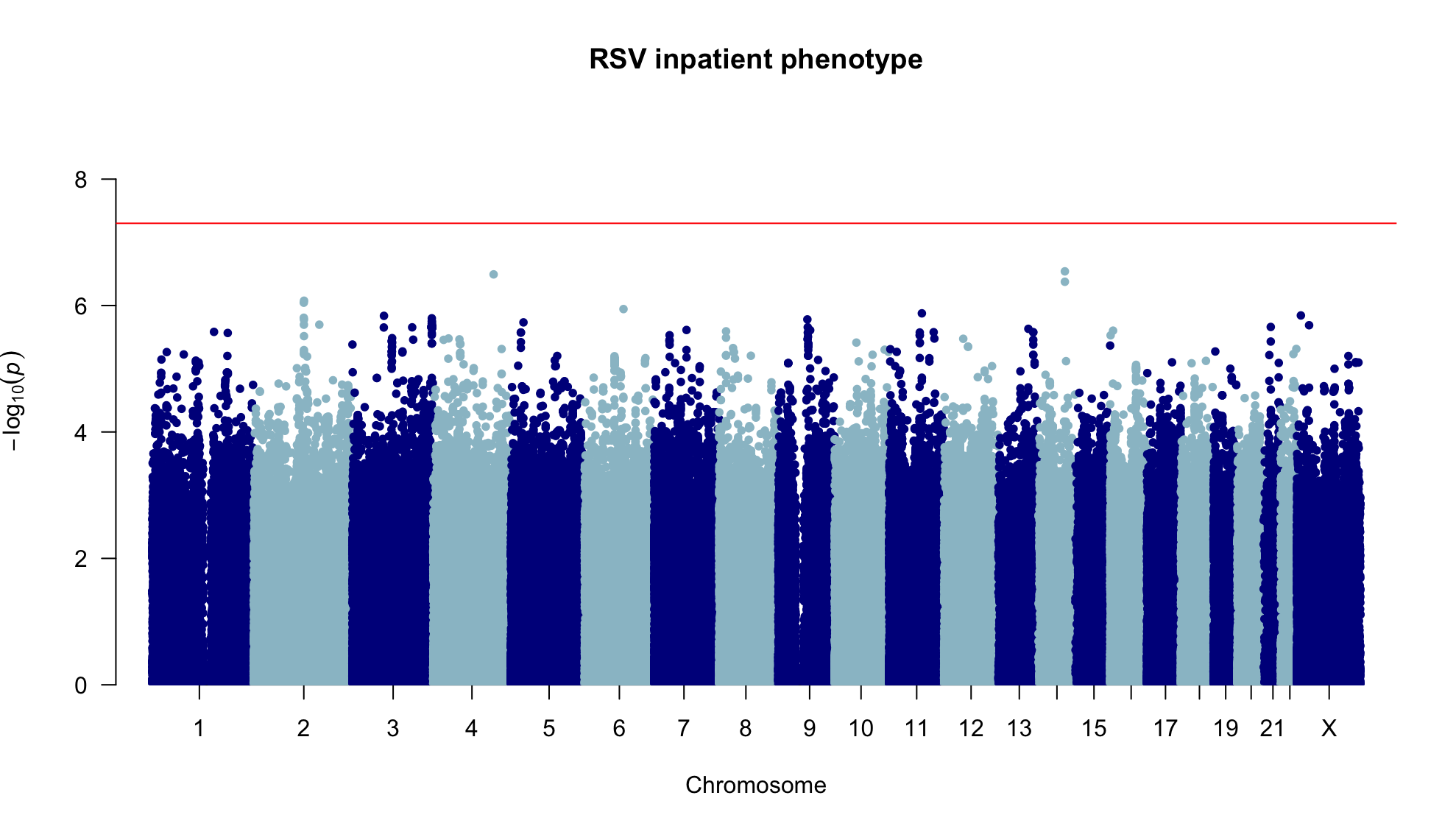

**Supplementary figure 6** Manhattan plot showing the results of inpatient-only RSV genome-wide association analysis (GWAS). This analysis was run in the FinnGen cohort (in contrast to the main analysis being meta-analysed across participating cohorts). In this analysis, the number of inpatient RSV cases was 403 with 22 650 controls, compared to the 467 cases and 22 586 controls in the main GWAS phenotype of in- and outpatient cases. There were thus only 64 outpatient cases in FinnGen. The genetic correlation between the inpatient phenotype and in-/outpatient phenotype was practically equal to one (rg = 1.05, SE = 0.07, p = 2.9×10^-52^), ie they showed virtually identical genome-wide signal. The lead meta-analysis variant rs787036 showed the same direction and similar effect size in the main in-/outpatient phenotype (beta = 0.287, p = 0.0016) and the inpatient-only sensitivity phenotype (beta = 0.259, p = 0.013).

We hypothesise that this highly overlapping genetic signal between the two phenotypes is due to the incomplete recording and ascertainment of milder RSV cases.

#### S7: Two-sample mendelian randomisation (MR) on genetic asthma liability to RSV susceptibility

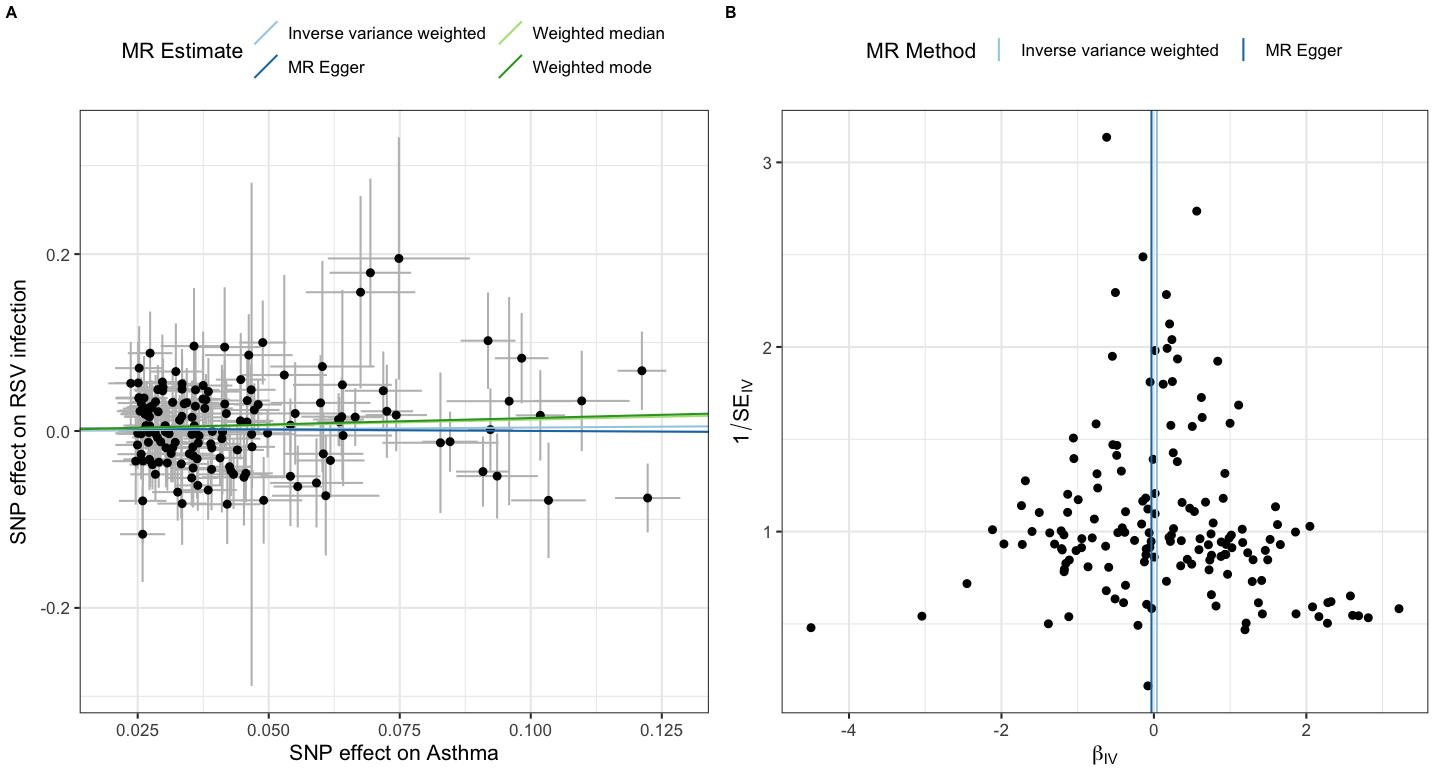

**Supplementary Figure 7** In the Two-Sample Mendelian Randomisation (MR) analysis, we selected 155 LD-independent (r²<0.001) genome-wide significant single nucleotide polymorphisms (SNP) for asthma (we excluded 1 SNP due to incompatible alleles and 4 due to palindromic SNP labelling and intermediate allele frequencies). The primary inverse-variance–weighted analysis showed no evidence of a causal effect (OR per log-odds increase in asthma liability 1.04, 95% CI 0.91–1.19; p = 0.58). Sensitivity analyses using MR-Egger (OR 0.97, 95% CI 0.69–1.35; p = 0.85), weighted median (OR 1.14, 95% CI 0.93–1.39; p = 0.21) and weighted mode (OR 1.16, 95% CI 0.77–1.73; p = 0.48) yielded similarly null estimates; there was no evidence of directional pleiotropy (p = 0.64), low heterogeneity (Q(p) = 0.39), and strong instruments (mean F value = 74). Leave-one-out analyses did not suggest that the estimate was driven by any single variant (data not shown). SNPs used in this analysis are listed in ***Supplementary Table 5.***

**Panel A)** Scatter plot of SNP-specific associations with asthma (x-axis, log-odds scale) and RSV infection (y-axis). Each point represents one asthma-associated SNP; grey bars show the corresponding standard errors. Coloured lines indicate the fitted MR estimates from the MR methods, all lying close to a null slope.

**Panel B)** Funnel plot of individual SNP ratio estimates (β_IV, x-axis) against their precision (1/SE_IV, y-axis). Points symmetrically scattered around the vertical lines representing the inverse-variance–weighted and MR-Egger estimates suggest no strong evidence of directional pleiotropy.
