## Supplementary material for "Infant respiratory syncytial virus and childhood asthma: a nationwide registry-based, sibling-controlled, and genome-wide association study": STROBE-MR

**STROBE-MR checklist of recommended items to address in reports of Mendelian randomization studies**^1^ ^2^

| **Item No.** | **Section** | **Checklist item** | **Page No.** | **Comments** |
| --- | --- | --- | --- | --- |
| 1 | **TITLE and ABSTRACT** | Indicate Mendelian randomization (MR) as the study’s design in the title and/or the abstract if that is a main purpose of the study | Abstract | Not in title, not the main purpose |
|  | **INTRODUCTION** |  |  |  |
| 2 | **Background** | Explain the scientific background and rationale for the reported study. What is the exposure? Is a potential causal relationship between exposure and outcome plausible? Justify why MR is a helpful method to address the study question | p2 | MR helpful to assess asthma -> RSV, because asthma genetic risk is well known. |
| 3 | **Objectives** | State specific objectives clearly, including pre-specified causal hypotheses (if any). State that MR is a method that, under specific assumptions, intends to estimate causal effects | p2-3 |  |
|  | **METHODS** |  |  |  |
| 4 | **Study design and data sources** | Present key elements of the study design early in the article. Consider including a table listing sources of data for all phases of the study. For each data source contributing to the analysis, describe the following: | p3 |  |
|  | a) | Setting: Describe the study design and the underlying population, if possible. Describe the setting, locations, and relevant dates, including periods of recruitment, exposure, follow-up, and data collection, when available. | p3, p5  suppl p4-5 | Both genotyped cohorts and registry-based cohorts described |
|  | b) | Participants: Give the eligibility criteria, and the sources and methods of selection of participants. Report the sample size, and whether any power or sample size calculations were carried out prior to the main analysis | p3, p5  suppl p4-5 |  |
|  | c) | Describe measurement, quality control and selection of genetic variants | Suppl p4-5;  Suppl p9 |  |
|  | d) | For each exposure, outcome, and other relevant variables, describe methods of assessment and diagnostic criteria for diseases | P3-5, p7; Suppl p6-7 |  |
|  | e) | Provide details of ethics committee approval and participant informed consent, if relevant | suppl P2-6 |  |
| 5 | **Assumptions** | Explicitly state the three core IV assumptions for the main analysis (relevance, independence and exclusion restriction) as well assumptions for any additional or sensitivity analysis | Suppl P9-10 |  |
| 6 | **Statistical methods: main analysis** | Describe statistical methods and statistics used | P6-7;  Suppl p6-10 |  |
|  | a) | Describe how quantitative variables were handled in the analyses (i.e., scale, units, model) | p6-7 |  |
|  | b) | Describe how genetic variants were handled in the analyses and, if applicable, how their weights were selected | Suppl P9-10 |  |
|  | c) | Describe the MR estimator (e.g. two-stage least squares, Wald ratio) and related statistics. Detail the included covariates and, in case of two-sample MR, whether the same covariate set was used for adjustment in the two samples | Suppl p9 | Covariate sets were different, as the Asthma GWAS was adjusted with the meta-analysis population covariates; we used our cohort-specific covariates. |
|  | d) | Explain how missing data were addressed | p6;  suppl p9-10 |  |
|  | e) | If applicable, indicate how multiple testing was addressed | Suppl p8 | Applicaple to discovery GWAS only |
| 7 | **Assessment of assumptions** | Describe any methods or prior knowledge used to assess the assumptions or justify their validity | Suppl p9 | Very little known about the outcome (RSV) genetic risk factors, also not many findings in our discovery GWAS; risk factors for severe RSV not likely to have genetic associations shared with asthma. |
| 8 | **Sensitivity analyses and additional analyses** | Describe any sensitivity analyses or additional analyses performed (e.g. comparison of effect estimates from different approaches, independent replication, bias analytic techniques, validation of instruments, simulations) | Suppl p9 | in the context of MR, sensitivity analysis was done using only the childhood asthma -associated variants. |
| 9 | **Software and pre-registration** |  |  |  |
|  | a) | Name statistical software and package(s), including version and settings used | p6;  suppl p9 |  |
|  | b) | State whether the study protocol and details were pre-registered (as well as when and where) | p5 | Study protocol not registered; GWAS analysis plan formalised and shared between consortium members |
|  | **RESULTS** |  |  |  |
| 10 | **Descriptive data** |  |  |  |
|  | a) | Report the numbers of individuals at each stage of included studies and reasons for exclusion. Consider use of a flow diagram | Figure 1; p13 | Flow diagram applicable to registry-based analyses only. |
|  | b) | Report summary statistics for phenotypic exposure(s), outcome(s), and other relevant variables (e.g. means, SDs, proportions) | p9-12 (registry analyses) p13-15 (genetic analyses) |  |
|  | c) | If the data sources include meta-analyses of previous studies, provide the assessments of heterogeneity across these studies | p13, Figure 5b, Supplementary table 4 |  |
|  | d) | For two-sample MR:  i.  Provide justification of the similarity of the genetic variant-exposure associations between the exposure and outcome samples  ii.  Provide information on the number of individuals who overlap between the exposure and outcome studies | Suppl p9-10 | We did not use individual-level data from the asthma GWAS and could not study quantify sample overlap between the published asthma exposure GWAS and the RSV outcome GWAS. However, the asthma exposure GWAS was very large with >150 000 cases across 22 biobanks, and any possible overlap is expected to be very limited, and the MR was performed using strong external asthma instruments. |
| 11 | **Main results** |  |  |  |
|  | a) | Report the associations between genetic variant and exposure, and between genetic variant and outcome, preferably on an interpretable scale | Supplementary table S5 & S6 |  |
|  | b) | Report MR estimates of the relationship between exposure and outcome, and the measures of uncertainty from the MR analysis, on an interpretable scale, such as odds ratio or relative risk per SD difference | Supplementary figure 7; supplementary table 7 |  |
|  | c) | If relevant, consider translating estimates of relative risk into absolute risk for a meaningful time period |  | not relevant in the current paper |
|  | d) | Consider plots to visualize results (e.g. forest plot, scatterplot of associations between genetic variants and outcome versus between genetic variants and exposure) | Supplementary figure 7 |  |
| 12 | **Assessment of assumptions** |  |  |  |
|  | a) | Report the assessment of the validity of the assumptions | P15; Supplementary figure 7; Supplementary table 7 |  |
|  | b) | Report any additional statistics (e.g., assessments of heterogeneity across genetic variants, such as *I^2^*, Q statistic or E-value) | P15; Supplementary figure 7; Supplementary table 7 |  |
| 13 | **Sensitivity analyses and additional analyses** |  |  |  |
|  | a) | Report any sensitivity analyses to assess the robustness of the main results to violations of the assumptions | P15; Supplementary figure 7; Supplementary table 7 | In the MR context, our sensitivity analysis was to run MR only with childhood asthma -specific variants. |
|  | b) | Report results from other sensitivity analyses or additional analyses |  |  |
|  | c) | Report any assessment of direction of causal relationship (e.g., bidirectional MR) | p15 | The other direction (RSV -> asthma) was not possible to analyse |
|  | d) | When relevant, report and compare with estimates from non-MR analyses |  | Not relevant here |
|  | e) | Consider additional plots to visualize results (e.g., leave-one-out analyses) | Supplementary figure 7 | Supplementary figure 7 shows variant associations; leave-one-out analyses are mentioned but data not shown. |
|  | **DISCUSSION** |  |  |  |
| 14 | **Key results** | Summarize key results with reference to study objectives | p15-16 |  |
| 15 | **Limitations** | Discuss limitations of the study, taking into account the validity of the IV assumptions, other sources of potential bias, and imprecision. Discuss both direction and magnitude of any potential bias and any efforts to address them | p17-18 |  |
| 16 | **Interpretation** |  |  |  |
|  | a) | Meaning: Give a cautious overall interpretation of results in the context of their limitations and in comparison with other studies | p17-18 |  |
|  | b) | Mechanism: Discuss underlying biological mechanisms that could drive a potential causal relationship between the investigated exposure and the outcome, and whether the gene-environment equivalence assumption is reasonable. Use causal language carefully, clarifying that IV estimates may provide causal effects only under certain assumptions | P16, P17 | Mechanisms discussed only cautiously. |
|  | c) | Clinical relevance: Discuss whether the results have clinical or public policy relevance, and to what extent they inform effect sizes of possible interventions | P17, P18 |  |
| 17 | **Generalizability** | Discuss the generalizability of the study results (a) to other populations, (b) across other exposure periods/timings, and (c) across other levels of exposure | P17-P18 (limitations) |  |
|  | **OTHER INFORMATION** |  |  |  |
| 18 | **Funding** | Describe sources of funding and the role of funders in the present study and, if applicable, sources of funding for the databases and original study or studies on which the present study is based | Abstract; p18-19 |  |
| 19 | **Data and data sharing** | Provide the data used to perform all analyses or report where and how the data can be accessed, and reference these sources in the article. Provide the statistical code needed to reproduce the results in the article, or report whether the code is publicly accessible and if so, where | p18 |  |
| 20 | **Conflicts of Interest** | All authors should declare all potential conflicts of interest | P19 |  |

This checklist is copyrighted by the Equator Network under the Creative Commons Attribution 3.0 Unported (CC BY 3.0) license.

1.	Skrivankova VW, Richmond RC, Woolf BAR, Yarmolinsky J, Davies NM, Swanson SA, et al. Strengthening the Reporting of Observational Studies in Epidemiology using Mendelian Randomization (STROBE-MR) Statement. JAMA. 2021;under review.

2.	Skrivankova VW, Richmond RC, Woolf BAR, Davies NM, Swanson SA, VanderWeele TJ, et al. Strengthening the Reporting of Observational Studies in Epidemiology using Mendelian Randomisation (STROBE-MR): Explanation and Elaboration. BMJ. 2021;375:n2233.
