## Supplementary material for "Infant respiratory syncytial virus and childhood asthma: a nationwide registry-based, sibling-controlled, and genome-wide association study": STREGA

### Reporting checklist for genetic association study.

Based on the STREGA guidelines.

#### Instructions to authors

Complete this checklist by entering the page numbers from your manuscript where readers will find each of the items listed below.

Your article may not currently address all the items on the checklist. Please modify your text to include the missing information. If you are certain that an item does not apply, please write "n/a" and provide a short explanation.

Upload your completed checklist as an extra file when you submit to a journal.

In your methods section, say that you used the STREGAreporting guidelines, and cite them as:

Little J, Higgins JP, Ioannidis JP, Moher D, Gagnon F, von Elm E, Khoury MJ, Cohen B, Davey-Smith G, Grimshaw J, Scheet P, Gwinn M, Williamson RE, Zou GY, Hutchings K, Johnson CY, Tait V, Wiens M, Golding J, van Duijn C, McLaughlin J, Paterson A, Wells G, Fortier I, Freedman M, Zecevic M, King R, Infante-Rivard C, Stewart A, Birkett N; STrengthening the REporting of Genetic Association Studies. STrengthening the REporting of Genetic Association Studies (STREGA): An Extension of the STROBE Statement.

|  |  | Reporting Item | Page Number | Additional remarks |
| --- | --- | --- | --- | --- |
| **Title and abstract** |  |  |  |  |
| Title | [#1a](https://www.goodreports.org/reporting-checklists/strega/info/#1a) | Indicate the study’s design with a commonly used term in the title or the abstract | [title] |  |
| Abstract | [#1b](https://www.goodreports.org/reporting-checklists/strega/info/#1b) | Provide in the abstract an informative and balanced summary of what was done and what was found | [abstract] |  |
| **Background/rationale** |  |  |  |  |
|  | [#2](https://www.goodreports.org/reporting-checklists/strega/info/#2) | Explain the scientific background and rationale for the investigation being reported | 3 |  |
| **Objectives** |  |  |  |  |
|  | [#3](https://www.goodreports.org/reporting-checklists/strega/info/#3) | State specific objectives, including any prespecified hypotheses. State if the study is the first report of a genetic association, a replication effort, or both. | p3 | both, stated in methods |
| **Study design** |  |  |  |  |
|  | [#4](https://www.goodreports.org/reporting-checklists/strega/info/#4) | Present key elements of study design early in the paper | p4 | Subheader “Study Design” |
| **Setting** |  |  |  |  |
|  | [#5](https://www.goodreports.org/reporting-checklists/strega/info/#5) | Describe the setting, locations, and relevant dates, including periods of recruitment, exposure, follow-up, and data collection | p4-6; Suppl p2-5 | Described. |
| **Eligibility criteria** |  |  |  |  |
|  | [#6a](https://www.goodreports.org/reporting-checklists/strega/info/#6a) | Cohort study – Give the eligibility criteria, and the sources and methods of selection of participants. Describe methods of follow-up. Case-control study – Give the eligibility criteria, and the sources and methods of case ascertainment and control selection. Give the rationale for the choice of cases and controls. Cross-sectional study – Give the eligibility criteria, and the sources and methods of selection of participants. Give information on the criteria and methods for selection of subsets of participants from a larger study, when relevant. | p4-8; Figure 1; Suppl p2-8 | Rationale in Methods – p6 and p7 |
|  | [#6b](https://www.goodreports.org/reporting-checklists/strega/info/#6b) | Cohort study – For matched studies, give matching criteria and number of exposed and unexposed. Case-control study – For matched studies, give matching criteria and the number of controls per case. | N/A |  |
| **Variables** |  |  |  |  |
|  | [#7a](https://www.goodreports.org/reporting-checklists/strega/info/#7a) | Clearly define all outcomes, exposures, predictors, potential confounders, and effect modifiers. Give diagnostic criteria, if applicable | p4-6, Supplements p6 |  |
|  | [#7b](https://www.goodreports.org/reporting-checklists/strega/info/#7b) | Clearly define genetic exposures (genetic variants) using a widely-used nomenclature system. Identify variables likely to be associated with population stratification (confounding by ethnic origin). | p7-8; Supplements p7-8. |  |
| **Data sources/measurement** |  |  |  |  |
|  | [#8a](https://www.goodreports.org/reporting-checklists/strega/info/#8a) | For each variable of interest give sources of data and details of methods of assessment (measurement). Describe comparability of assessment methods if there is more than one group. Give information separately for for exposed and unexposed groups if applicable. | Methods p4-6; Supplements p2-8 | Generally, all data is collected from routinely maintained nationwide registries. |
|  | [#8b](https://www.goodreports.org/reporting-checklists/strega/info/#8b) | Describe laboratory methods, including source and storage of DNA, genotyping methods and platforms (including the allele calling algorithm used, and its version), error rates and call rates. State the laboratory / centre where genotyping was done. Describe comparability of laboratory methods if there is more than one group. Specify whether genotypes were assigned using all of the data from the study simultaneously or in smaller batches. | Supplements p4-6 | General description of cohrots and given in supplements. As this is a meta-analytic study combining data from multiple cohorts with established pipelines cohort publications, we refer to original publications for details. |
| **Bias** |  |  |  |  |
|  | [#9a](https://www.goodreports.org/reporting-checklists/strega/info/#9a) | Describe any efforts to address potential sources of bias | p4, p8-10 | Also discussed in ‘strenghts and limitations’ |
|  | [#9b](https://www.goodreports.org/reporting-checklists/strega/info/#9b) | Describe any efforts to address potential sources of bias |  |  |
| **Study size** |  |  |  |  |
|  | [#10](https://www.goodreports.org/reporting-checklists/strega/info/#10) | Explain how the study size was arrived at | N/A | In an attempt to do a largest GWAS, all available and suitable data were used. Similarly in registry analyses, we used all available data. |
| **Quantitative variables** |  |  |  |  |
|  | [#11](https://www.goodreports.org/reporting-checklists/strega/info/#11) | Explain how quantitative variables were handled in the analyses. If applicable, describe which groupings were chosen, and why. If applicable, describe how effects of treatment were dealt with. | p6-7 | All available continuous variables were treated as continuous. In the case of continuous covariates (birth year, gestational age) they were modelled as linear. |
| **Statistical methods** |  |  |  |  |
|  | [#12a](https://www.goodreports.org/reporting-checklists/strega/info/#12a) | Describe all statistical methods, including those used to control for confounding. State software version used and options (or settings) chosen. | p6-7; Supplements p7-9 | Specific R packages also mentioned in Supplementary Figure and Supplementary Table legends |
|  | [#12b](https://www.goodreports.org/reporting-checklists/strega/info/#12b) | Describe any methods used to examine subgroups and interactions | p6 |  |
|  | [#12c](https://www.goodreports.org/reporting-checklists/strega/info/#12c) | Explain how missing data were addressed | p7; supplements p3-4 | Missingness was rare and absence of event is considered as no event in routine registries. In the case of missing data, those individuals were excluded from analyses |
|  | [#12d](https://www.goodreports.org/reporting-checklists/strega/info/#12d) | If applicable, explain how loss to follow-up was addressed | p7; supplements p3-4 |  |
|  | [#12e](https://www.goodreports.org/reporting-checklists/strega/info/#12e) | Describe any sensitivity analyses | p6, p8; suppl p10 |  |
|  | [#12f](https://www.goodreports.org/reporting-checklists/strega/info/#12f) | State whether Hardy-Weinberg equilibrium was considered and, if so, how. | p7; Suppl p7 |  |
|  | [#12g](https://www.goodreports.org/reporting-checklists/strega/info/#12g) | Describe any methods used for inferring genotypes or haplotypes | Supplements p3-5, p7 | For details of imputation in original participating studies, we refer to cohort descriptions |
|  | [#12h](https://www.goodreports.org/reporting-checklists/strega/info/#12h) | Describe any methods used to assess or address population stratification. | p6, Supplements p8 |  |
|  | [#12i](https://www.goodreports.org/reporting-checklists/strega/info/#12i) | Describe any methods used to address multiple comparisons or to control risk of false positive findings. | p6-7, Supplements p8 |  |
|  | [#12j](https://www.goodreports.org/reporting-checklists/strega/info/#12j) | Describe any methods used to address and correct for relatedness among subjects | Suppl p8 | Recommended to use mixed model software, as standard. |
| **Participants** |  |  |  |  |
|  | [#13a](https://www.goodreports.org/reporting-checklists/strega/info/#13a) | Report numbers of individuals at each stage of study—eg numbers potentially eligible, examined for eligibility, confirmed eligible, included in the study, completing follow-up, and analysed. Give information separately for for exposed and unexposed groups if applicable. Report numbers of individuals in whom genotyping was attempted and numbers of individuals in whom genotyping was successful. | Figure 1; Suppl p 5-6, Supplementary table 3 | Related to GWAS meta-analysis, data was provided from established existing cohorts and two previous publication. We have not reproduced numbers of overall genotyped subjects, and related to previous publications, we don’t have access to that data. In population-based cohorts and replication cohort, detailed information for each step of exclusions are given. Number of successfully genotyped persons is described as part of original cohort descriptions. |
|  | [#13b](https://www.goodreports.org/reporting-checklists/strega/info/#13b) | Give reasons for non-participation at each stage | Figure 1, p4 and p6, Suppl p5-6 | Given for population level cohorts and replication cohort MoBa, not for GWAS meta-analysis cohorts, information elsewhere described. |
|  | [#13c](https://www.goodreports.org/reporting-checklists/strega/info/#13c) | Consider use of a flow diagram | Figure 1 | Figure 1 for population-level cohorts, not for genotyped cohorts |
| **Descriptive data** |  |  |  |  |
|  | [#14a](https://www.goodreports.org/reporting-checklists/strega/info/#14a) | Give characteristics of study participants (eg demographic, clinical, social) and information on exposures and potential confounders. Give information separately for exposed and unexposed groups if applicable. Consider giving information by genotype | Supplementary table 1 | Not given for exposure status (RSV hospitalization) or for genotype. |
|  | [#14b](https://www.goodreports.org/reporting-checklists/strega/info/#14b) | Indicate number of participants with missing data for each variable of interest | p7; supplements p3, p6 | Listed, and also discussed; generally missingness was rare. |
|  | [#14c](https://www.goodreports.org/reporting-checklists/strega/info/#14c) | Cohort study – Summarize follow-up time, e.g. average and total amount. | p4-5, p7 |  |
| **Outcome data** |  |  |  |  |
|  | [#15](https://www.goodreports.org/reporting-checklists/strega/info/#15) | Cohort study Report numbers of outcome events or summary measures over time. Give information separately for exposed and unexposed groups if applicable. Report outcomes (phenotypes) for each genotype category over time Case-control study – Report numbers in each exposure category, or summary measures of exposure.Give information separately for cases and controls . Report numbers in each genotype category. Cross-sectional study – Report numbers of outcome events or summary measures. Give information separately for exposed and unexposed groups if applicable. Report outcomes (phenotypes) for each genotype category | p10; p12 (figure 3 text) | Individual genotype counts were not available for the meta-analysis of summary statistics, but we provide the total sample size, allele frequencies (AF) and AF variance, effect size and direction of effect with CI’s in each cohort for lead SNPs. Quality control parameters (including HWE P-value thresholds and genotyping call rates) were imposed in our analysis protocol, and described in methods, and no deviations were reported. The lead locus (rs787036) passed all standard quality control filters in the original discovery cohorts |
| **Main results** |  |  |  |  |
|  | [#16a](https://www.goodreports.org/reporting-checklists/strega/info/#16a) | Give unadjusted estimates and, if applicable, confounder-adjusted estimates and their precision (eg, 95% confidence interval). Make clear which confounders were adjusted for and why they were included | p10, p13, Supplementary table 2 | In the sibling-pair conditional model, coefficients from only the covariate-adjusted model are given, but we report the crude unadjusted risk ratio in the population. |
|  | [#16b](https://www.goodreports.org/reporting-checklists/strega/info/#16b) | Report category boundaries when continuous variables were categorized | Supplementary table 1 | No categorization for other than descriptive purposes in background characteristics (supplementary table 1) |
|  | [#16c](https://www.goodreports.org/reporting-checklists/strega/info/#16c) | If relevant, consider translating estimates of relative risk into absolute risk for a meaningful time period | N/A |  |
|  | [#16d](https://www.goodreports.org/reporting-checklists/strega/info/#16d) | Report results of any adjustments for multiple comparisons | Supplements p8 | In GWAS, threshold for multiple-testing corrected statistical significance is generally 5e-8 |
| **Other analyses** |  |  |  |  |
|  | [#17a](https://www.goodreports.org/reporting-checklists/strega/info/#17a) | Report other analyses done—e.g., analyses of subgroups and interactions, and sensitivity analyses | p8, supplements p9-10, The whole results section | No interaction analyses. Sensitivity analyses included GAMM (Supplementary Figure 1) and MR with COA-specific SNPs (Supplementary table 7) |
|  | [#17b](https://www.goodreports.org/reporting-checklists/strega/info/#17b) | Report other analyses done—e.g., analyses of subgroups and interactions, and sensitivity analyses |  |  |
|  | [#17c](https://www.goodreports.org/reporting-checklists/strega/info/#17c) | Report other analyses done—e.g., analyses of subgroups and interactions, and sensitivity analyses |  |  |
| **Key results** |  |  |  |  |
|  | [#18](https://www.goodreports.org/reporting-checklists/strega/info/#18) | Summarise key results with reference to study objectives | p16 (discussion) |  |
| **Limitations** |  |  |  |  |
|  | [#19](https://www.goodreports.org/reporting-checklists/strega/info/#19) | Discuss limitations of the study, taking into account sources of potential bias or imprecision. Discuss both direction and magnitude of any potential bias. | p19 |  |
| **Interpretation** |  |  |  |  |
|  | [#20](https://www.goodreports.org/reporting-checklists/strega/info/#20) | Give a cautious overall interpretation considering objectives, limitations, multiplicity of analyses, results from similar studies, and other relevant evidence. | p16-18 |  |
| **Generalisability** |  |  |  |  |
|  | [#21](https://www.goodreports.org/reporting-checklists/strega/info/#21) | Discuss the generalisability (external validity) of the study results | p16-18, in particular p18 |  |
| **Funding** |  |  |  |  |
|  | [#22](https://www.goodreports.org/reporting-checklists/strega/info/#22) | Give the source of funding and the role of the funders for the present study and, if applicable, for the original study on which the present article is based | Abstract, Aknowledgements |  |

None The STREGA checklist is distributed under the terms of the Creative Commons Attribution License CC-BY. This checklist can be completed online using <https://www.goodreports.org/>, a tool made by the [EQUATOR Network](https://www.equator-network.org) in collaboration with [Penelope.ai](https://www.penelope.ai)
